# Home working and its association with social and mental wellbeing at different stages of the COVID-19 pandemic: Evidence from seven UK longitudinal population surveys

**DOI:** 10.1101/2022.10.03.22280412

**Authors:** Jacques Wels, Bożena Wielgoszewska, Bettina Moltrecht, Charlotte Booth, Michael J Green, Olivia KL Hamilton, Evangelia Demou, Giorgio Di Gessa, Charlotte Huggins, Jingmin Zhu, Gillian Santorelli, Richard J. Silverwood, Daniel Kopasker, Richard J. Shaw, Alun Hughes, Praveetha Patalay, Claire Steves, Nish Chaturvedi, David Porteous, Rebecca Rhead, Srinivasa Vittal Katikireddi, George B. Ploubidis

## Abstract

**Background:** Home working rates have increased since the COVID-19 pandemic’s onset, but the health implications of this transformation are unclear. We assessed the association between home working and social and mental wellbeing through harmonised analyses of seven UK longitudinal studies.

**Methods:** We estimated associations between home working and measures of psychological distress, low life satisfaction, poor self-rated health, low social contact, and loneliness across three different stages of the COVID-19 pandemic (T1= Apr-Jun 2020 – first lockdown, T2=Jul-Oct 2020 – eased restrictions, T3=Nov 2020-Mar 2021 – second lockdown), in seven population-based cohort studies using modified Poisson regression and meta-analyses to pool results across studies.

**Findings:** Among 34,131 observations spread over three time points, we found higher rates of home working at T1 and T3 compared with T2, reflecting lockdown periods. Home working was not associated with psychological distress at T1 (RR=0.92, 95%CI=0.79-1.08) or T2 (RR=0.99, 95%CI=0.88-1.11), but a detrimental association was found with psychological distress at T3 (RR=1.17, 95%CI=1.05-1.30). Poorer psychological distress associated with home working was observed for those educated to below degree level at T2 and T3. Men working from home reported poorer self-reported health at T2.

**Interpretation:** No clear evidence of an association between home working and mental wellbeing was found, apart from greater risk of psychological distress associated with home working during the second lockdown, but differences across sub-groups may exist. Longer term shifts to home working might not have adverse impacts on population wellbeing in the absence of pandemic restrictions but further monitoring of health inequalities is required.

**Funding:** National Core Studies, funded by UKRI, NIHR and the Health and Safety Executive.

## Introduction

Home working is rapidly increasing worldwide^1^. This shift had largely been voluntary until the COVID-19 pandemic, when home working became mandatory for many (but not all) workers. According to the International Labour Organisation, 557 million employees worldwide worked from home during the second quarter of 2020, accounting for 17 percent of the global workforce ^2^. In the UK, estimates were higher, with 37 percent of the workforce working from home in 2020 compared to the 27 to 30 percent who worked from home in 2019 ^3,4^. This sudden and widespread uptake in home working provides an opportunity to examine the potential impact of home working on the mental health and wellbeing of a diverse range of workers. This is of particular importance if higher levels of home working are sustained, as is expected.

The relationship between home working and mental health is poorly understood, with mixed pre-pandemic evidence and potential mechanisms for both positive ^5–7^ and negative impacts ^8,9^. Potential impacts of home working on health inequalities have been under-explored in the literature, so it is important to assess whether associations differ by social factors such as sex, age, education, and hours worked. The pandemic context also offers the opportunity to assess the relationship between home working and mental health under varying degrees of public restrictions, at different time points during the pandemic.

The UK National Core Studies Longitudinal Health and Wellbeing initiative draws together data from several UK population-based longitudinal studies, using coordinated analyses to answer pandemic-related questions. By conducting new harmonised analyses within each study and pooling estimates, we provide robust evidence on associations between home working and mental wellbeing during the pandemic, with a view to understanding the longer-term consequences of this shift. More specifically, we address the following two research questions (RQs): (RQ1) Is working from home (fully or partially) associated with psychological distress, low life satisfaction, low self-rated health, low social contact, and loneliness at different stages of the pandemic?; (RQ2) Do associations between home working and self-reported social and mental wellbeing differ by sex, age, education, or full-time versus part-time work?

## Methods

### Data, Design and Sample

We conducted primary data analyses in seven UK population-based studies, including three age-homogenous birth cohorts: Next Steps (NS, formerly the Longitudinal Study of Young People in England), the 1970 British Cohort Study (BCS70), and the 1958 National Child Development Study (NCDS); and four age heterogenous studies: Understanding Society – also referred to as the UK Household Longitudinal Study (USOC) –, the English Longitudinal Study of Ageing (ELSA), the Scottish Family Health Study – Generation Scotland (GS) –, and Born in Bradford (BiB). Details of all studies are presented in Supplementary file 1.

Participants were surveyed at multiple timepoints during three key periods of the pandemic (data availability by time point in Supplementary file 2). The first period (T1) included surveys from April-June 2020, during the initial surge of infections and the first national lockdown. The second period (T2) included surveys from July-October 2020, as initial restrictions were eased. The final period (T3) included surveys from November 2020-March 2021, as infection rates rose again, and a second national lockdown was introduced.

Analytical samples include respondents of working age aged 16 to 66 who were employed prior to the pandemic and actively employed (i.e., excluding those who were furloughed) during at least one of the pandemic time-points. The sample was restricted to complete cases and to respondents for whom information about mental health and social wellbeing was collected in both pandemic and pre-pandemic surveys.

All analyses were pre-planned in accordance with our published protocol ^10^.

### Measures

Measures and derived harmonised variables are described below, with further details on study-specific measurement in Supplementary file 3.

#### Exposure: Home working

Respondents in each study indicated whether they had been working from home fully, partially, or not at all, at each of the three pandemic timepoints. As NS, BCS70 and NCDS did not collect information about partial home working at T1, across all studies, we generated a “home working” variable with two modalities at T1 (0= “working entirely at employer’s premises or other location” ; 1=“Working fully or partially from home”) and three modalities at T2 and T3 (0= “working entirely at employer’s premise or other location”; 1= “partially working from home”; 2= “fully working from home”).

#### Outcomes: Social and mental wellbeing

We investigated five different outcomes: psychological distress, low life satisfaction, poor or fair self-rated health, low social contact (including online contacts), and loneliness. For each outcome, we created a binary indicator using pre-validated cut-off scores where possible (detailed information about measurement across studies in Supplementary file 3).

#### Covariates

Analyses for RQ1 were repeated with four nested levels of adjustment, with each including a progressively larger set of covariates variables:

1. No adjustment.
2. Socio-demographic adjustment: age (for age-heterogeneous studies; three categories: 16- 29, 30-49, 50+ that maximise population sizes by group), sex (male, female), housing tenure (mortgage or owner versus other), ethnicity (White -including White minorities- versus Ethnic Minority groups), level of education (university degree versus lower level of education) and household overcrowding (number of people in household/number of rooms). We also added a household composition variable with six categories, in line with the evidence suggesting women have been disproportionately burdened with childcare and home schooling during the pandemic ^11,12^ (‘alone’ (reference); ‘1=female with partner and child(ren)’; ‘2=male with partner and child(ren)’; ‘3=female with partner and no child(ren)’; ‘4=male with and partner no child(ren)’; ‘5=lone parent’; ‘6=others (i.e., living with other relatives or non-relatives)’).
3. Job adjustment: additionally included pre-pandemic weekly working hours, pre-pandemic Social Class (three class National Statistics Socio-Economic Classification; NS-SEC), 2-digit Standard Occupational Classification (SOC), and key worker status during the pandemic. We also controlled for propensity to be working from home prior to the start of the pandemic. Propensities were derived from an external source (the Annual Population Survey) ^13^ because no study except USOC collected information on pre-pandemic home working, These were calculated as the propensities to work fully or partially from home based on: SOC2010 (2- digits), sex, and age-group (16-29, 30-49 and 50-66) prior to the start of the pandemic (April 2019-March 2020), as described in Supplementary file 4.
4. Full adjustment: additionally controlled for pre-pandemic mental health, social wellbeing and the presence of a long-standing illness or disability, as described in Supplementary file 3. The fully adjusted model controls for pre-pandemic measures of the outcome and the resultant association can be interpreted as change in the outcomes compared to pre-pandemic levels.

Analyses for RQ2, which were stratified by sex, age, education level, and part-time versus full-time work used the full adjustment only.

### Analyses

For RQ1, we first ran within-study modified Poisson regression models with robust standard errors to model binary outcome variables ^14,15^ and report risk ratios (RR). RRs ease interpretation and avoid issues related to non-collapsibility of odds ratios ^16^. USOC had multiple surveys within each time period, so multi-level models were used to account for correlation between responses from the same individuals. We modelled the outcomes at each time point separately, using weights to account for survey non-response ^17^. Estimates from each model and study were then pooled using a random effects meta-analysis with restricted maximum likelihood.

Sensitivity analyses were conducted to validate the imputed propensity for pre-pandemic home working using data from USOC where actual reported pre-pandemic home working was available, to check for consistency with the imputed variable. Models for RQ1 were also repeated excluding data from GS and BiB (which did not have sufficient data for all levels of adjustment), to see whether results were consistent.

For RQ2, stratified analyses were conducted in the same way as above with subgroup meta-analyses performed by sex (male, female), age group (16-29, 30-49, 50+), education level (university degree, lower education level), and working hours (full-time, part-time) in order to assess between-group differences in the association between home working and social and mental wellbeing.

## Results

Across the seven longitudinal population studies, a total of 10,367 at T1, 11,585 at T2, and 12,179 at T3 individuals were included in the analyses. Before the start of the pandemic, 30.1% of the population reported working from home (data only available in USOC). This figure increased at T1 with percentages ranging between 32.9 and 65.5 across studies (Table 1). Percentages after combining working fully or partially from home were between 28.8 (in NCDS) and 64.7 (in BiB) percent at T2 and between 36.5 (in NCDS) to 64.2 (in GS) at T3. See Supplementary files 5 and 6 for a detailed overview of descriptive statistics for exposure, covariates, and outcome variables.

**Table 1.**
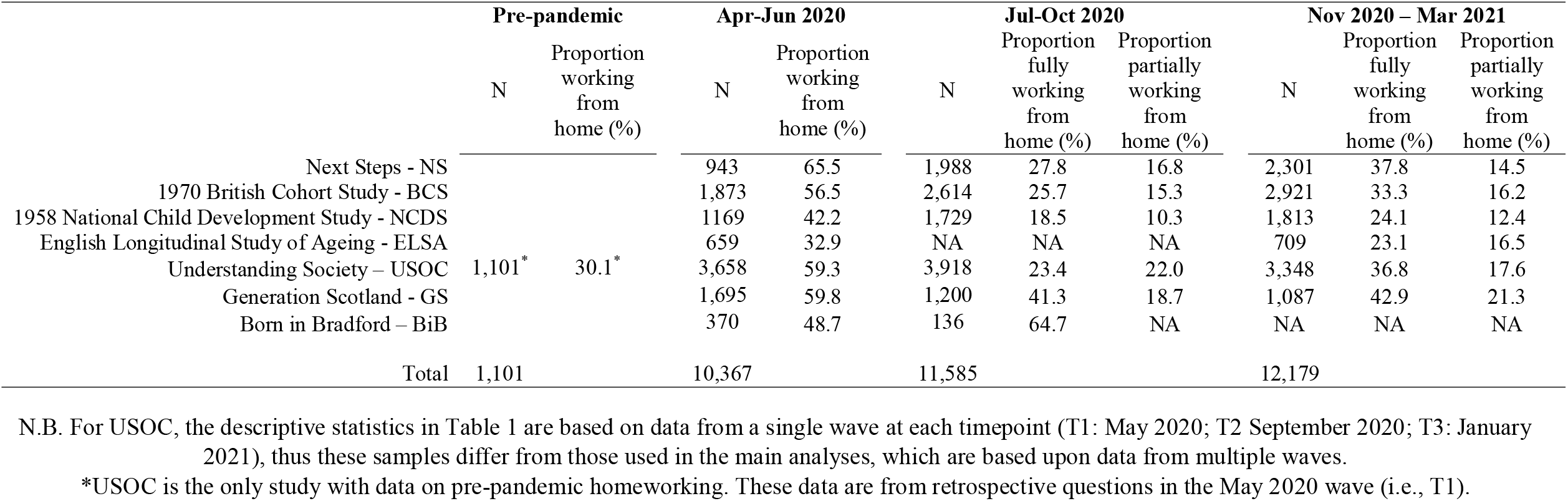
Home working by time point across the seven studies (weighted data)

Details of study-specific main estimates (RQ1) are presented in Supplementary file 7 with meta-analysed findings presented in Figure 1.

**Figure 1.**
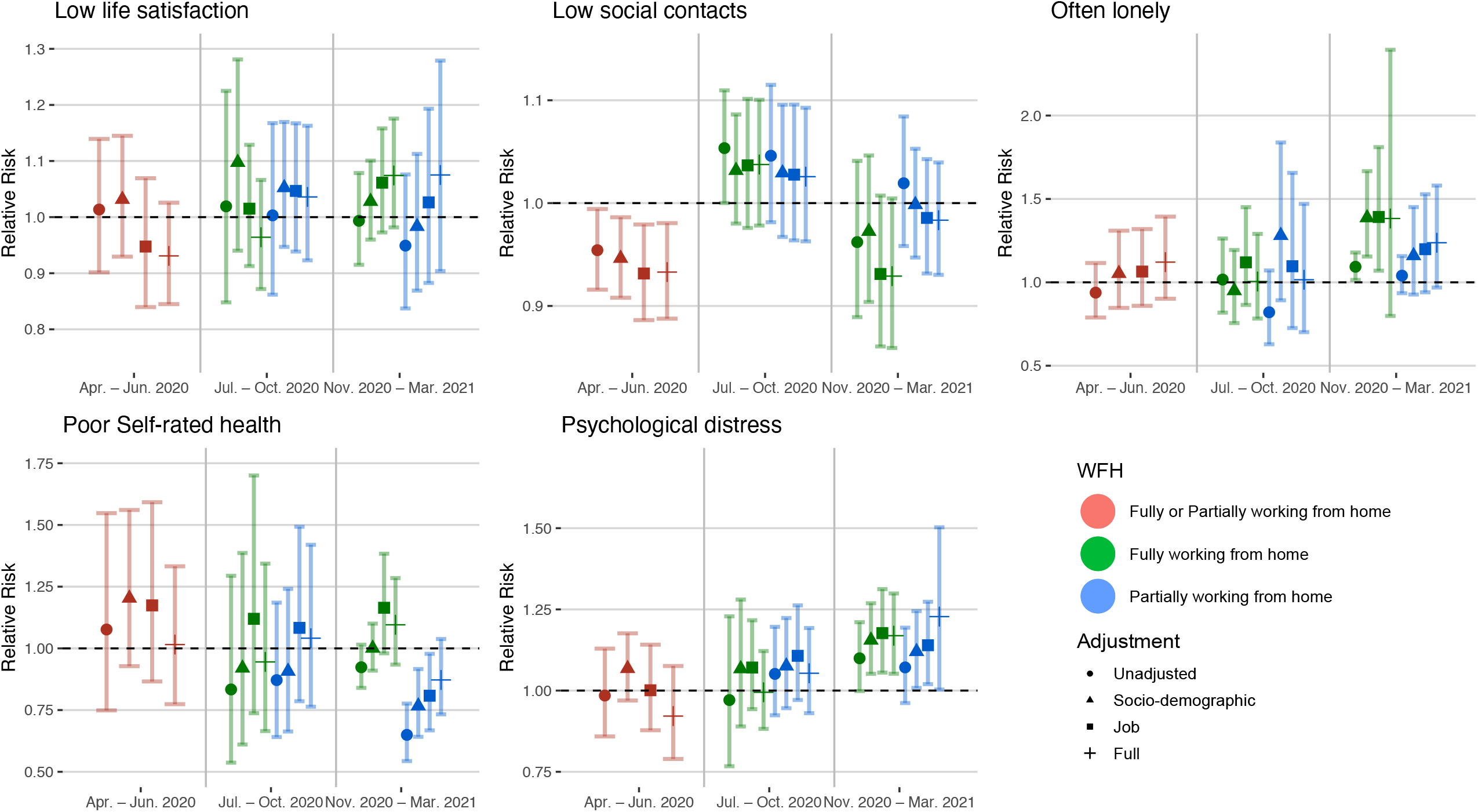
Main model estimates

There was no association between full or partial home working and low life satisfaction at T1, T2 or T3.

For social contact, working from home was associated with decreased risk of low social contact (fully adjusted RR: 0.93; 95% confidence interval (CI): 0.89-0.98) at T1. At T2, the association was attenuated for both fully and partially working from home, even reversing slightly (e.g., unadjusted RR for full home working: 1.05; 95% CI: 1.00-1.10). At T3, fully working from home was again associated with decreased risk of low social contact (fully adjusted RR: 0.93; 95% CI: 0.86-1.01), while the association for partial home working was still relatively attenuated (fully adjusted RR: 0.98; 95% CI: 0.93-1.04).

Regarding loneliness, there was little evidence for associations between home working and often feeling lonely at T1 and T2. At T3, fully working from home was associated with increased risk of often feeling lonely across the four levels of adjustment but associations were imprecisely estimated (fully adjusted RR: 1.38; 95% CI: 0.80-2.39). Partially working from home at T3 was similarly associated with often feeling lonely (fully adjusted RR: 1.24; 95% CI: 0.97-1.58).

For poor self-rated health, evidence did not support an association with full working from home at any time point. Partially working from home was associated with reduced risk of poor self-rated health at T3 (RR: 0.81; 95% CI: 0.67-0.98), but this was attenuated with adjustment (fully adjusted RR: 0.87; 95% CI: 0.73-1.04).

There was some indication of home working being associated with increased risk for psychological distress at T1 and T2, but CIs over-lapped the null. At T3, home working was associated with increased psychological distress for both fully working from home (fully adjusted RR: 1.17; 95% CI: 1.05-1.30) and partially working from home (fully adjusted RR: 1.22; 95% CI: 1.00-1.48).

Figures 2 and 3 show the estimates from the stratified meta-analyses (RQ2; full details of between-group heterogeneity tests are in Supplementary file 8). The observed associations between home working and social and mental wellbeing measures largely did not differ by sex, age, education, or full versus part-time work.

**Figure 2.**
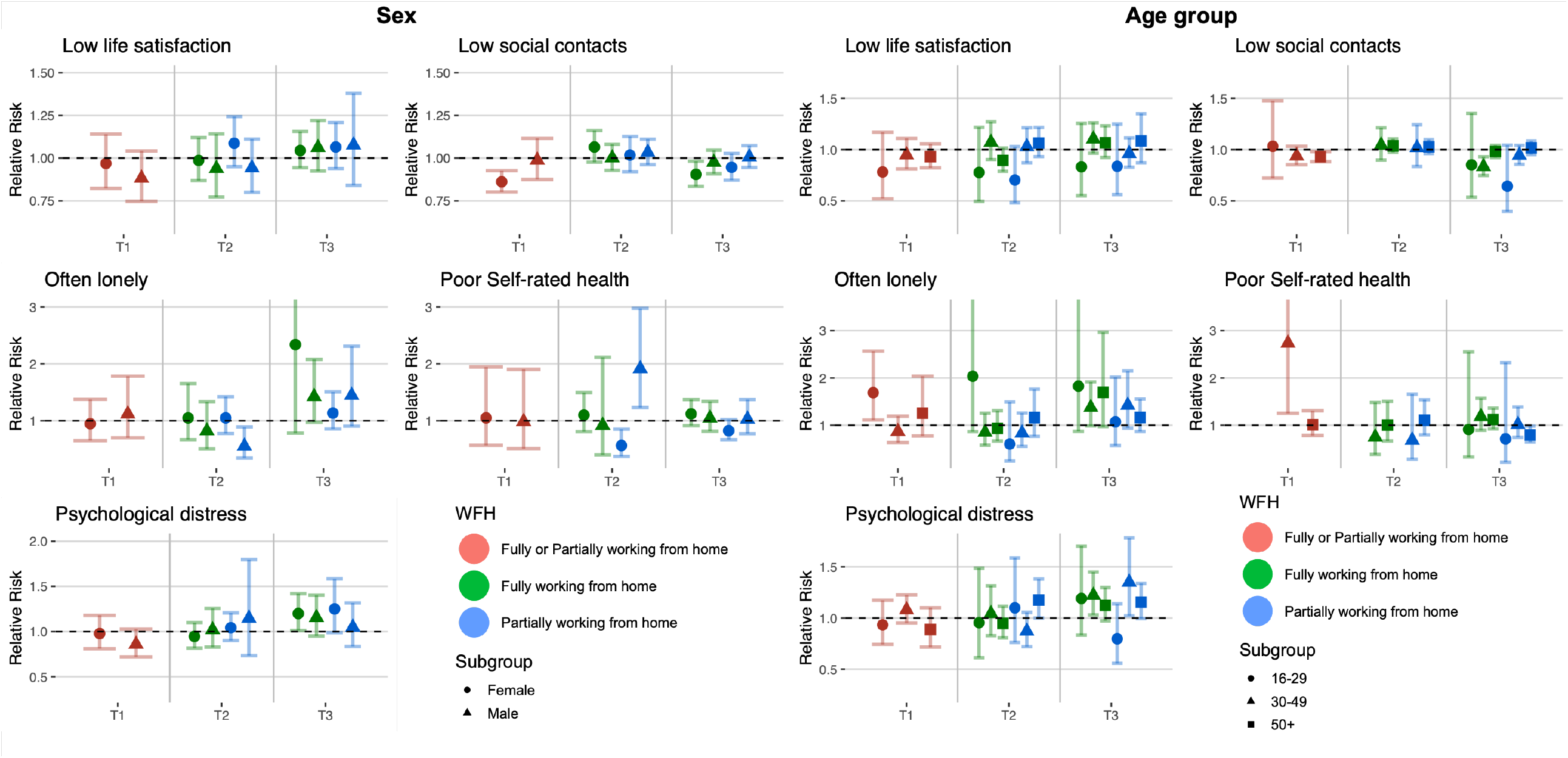
Stratified analyses by sex and age group

**Figure 3.**
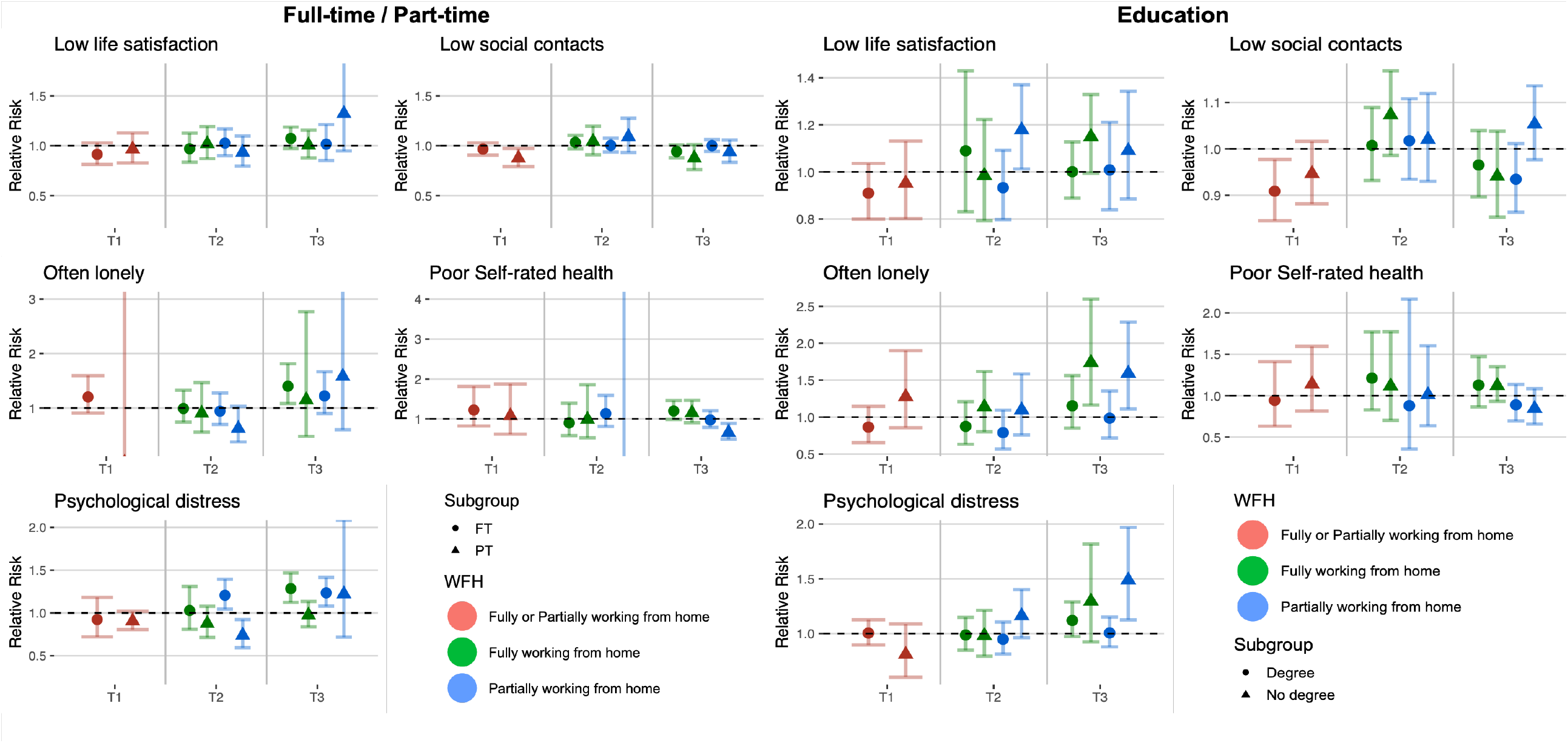
Stratified analyses by full-time and part-time distinction and education level

Looking at sex, associations between home working and reduced risk of low social contact appeared concentrated among females (T1 RR: 0.86; 95% CI: 0.80-0.93; T3 fully working from home RR: 0.91; 95% CI: 0.83-0.98) rather than males (T1 RR: 0.98; 95% CI: 0.87-1.11; T3 fully working from home RR: 0.97; 95% CI: 1.05-5.96). At T2, males who were partially working from home had reduced risk of often feeling lonely (RR: 0.55; 95% CI: 0.35-0.89) with no clear association among females (RR: 1.05; 95% CI: 0.78-1.42). Among those partially working from home, males showed increased risk of poor self-rated health (RR: 1.91; 95% CI: 1.23-2.98) in contrast to a reduced risk for females (RR: 0.57; 95% CI: 0.38-0.85).

Looking across age-groups, the association between fully working from home and reduced risk of low social contact at T3 was clearer in the 30-49 age group (RR: 0.83; 95% CI: 0.75- 0.93) than in the 50+ age group (RR: 0.98; 95% CI: 0.92-1.04). For the 16–29-year age group, there was evidence that home working was associated with increased risk of loneliness at T1 (RR: 1.69; 95% CI: 1.11-2.56). At T1, working from home was clearly associated with increased risk of poor self-rated health for respondents aged 30-49 (RR: 2.74; 95% CI: 1.25- 5.99), but not for those aged 50+ (RR: 1.01; 95% CI: 0.78-1.30), while no estimate was available for those aged 16-29 years.

Looking at part-time versus full-time work, partial home working was associated with increased risk of psychological distress for those working full-time at T2 (RR: 1.21; 95% CI: 1.05-1.39) but decreased risk for those working part-time (RR: 0.74; 95% CI: 0.59-0.92). Working fully from home was associated with increased psychological distress for those in full-time work at T3 (RR: 1.28; 95% CI: 1.12-1.47), but not for those in part-time work (RR: 0.98; 95% CI: 0.84-1.13).

Finally, looking at education, there was increased risk of low life satisfaction among respondents with no degree for partial home working at T2 (RR: 1.18; 95% CI: 1.01-1.37) and for full home working at T3 (RR: 1.15; 95% CI: 1.00-1.33) but not among participants with a degree. Furthermore, at T3 for both loneliness and psychological distress, there was a pattern whereby home working was more clearly associated with poor wellbeing among those with no degree (e.g., partial home working RR for distress: 1.49; 95% CI: 1.12-1.97) than those with a degree (RR: 1.01; 95% CI: 0.88-1.15).

Supplementary file 9 details re-analysis of the USOC data for the main model, using observed pre-pandemic home working (rather than the imputed values), showing consistent results. Additional analyses were also made excluding BiB and GS due to lack of consistency in the control and exposure variables with similar results observed (see Supplementary file 10) except for the association between loneliness and fully working from home that became significant at 95% at T3 (fully adjusted RR:1.52; 95% CI: 1.12-2.18) but with no change in coefficient intensity (RR:1.38; 95% CI: 0.80-2.39).

## Discussion

Analysing data from seven UK longitudinal population studies, with adjustment for a range of confounding factors, we found little supporting evidence for an association between home working and lower social and mental wellbeing during the first UK lockdown. Indeed, this study found only weak evidence that home working increased social contact during this period. There is little evidence of associations between home working and social and mental wellbeing when restrictions were eased during the summer of 2020.

The study also shows that some population sub-groups may have been affected. Partial home working was associated with increased risk of psychological distress for those aged 50 or over and for those working full-time. We also found an increased risk of poor self-rated health for males, and increased risk of low life satisfaction for those without degree-level education. When lockdown measures were re-introduced in the UK (late 2020 to early 2021), there was evidence that home working (full or partial) was associated with increased risk of loneliness and psychological distress across the population, especially for those aged 30-49 years, those without a degree, and those in full-time work. However, those apparent differences might be considered to just be noise as we do not find a consistent pattern across time-points and differences across sub-groups might require further monitoring.

Workers have experienced tremendous disruptions due to the pandemic ^18,19^, with many losing their job, being furloughed ^20^, experiencing changes in working hours ^21^, or shifting to working from home ^22,23^. The impact of employment disruptions on mental and social wellbeing during the pandemic has been investigated ^20^, but little is known about the role of home working. The clearest pattern of results emerged at T3, during a period of high infections and lockdown restrictions (between November 2020 – March 2021), when those working either partially or fully from home showed an increased risk for psychological distress and loneliness. At that time, the UK population was almost one year into the COVID- 19 pandemic, so the finding could represent people experiencing “lockdown fatigue” in relation to home working. As the pandemic progressed, people were also increasingly returning to work outside the home, so another explanation of these findings could be that those with poor mental wellbeing were more likely to maintain home working arrangements. Results appeared to be stronger in the age 30-49 bracket, for those without a degree, and for those working full-time. These demographic groups may have faced additional pressures on their time, due to childcare and home-schooling responsibilities. This finding shows that the relation between home working and social and mental wellbeing could be particularly sensitive to the overall pandemic context as well as the kind of work arrangement that is implemented. We also found a small decreased risk for low social contact among home workers during lockdowns. We speculate this could have been driven by more frequent social contact (e.g., via phone calls and messaging) which could have mitigated increased loneliness and poor mental health.

Pre-pandemic evidence suggests that home working is associated with multiple benefits, including greater employee productivity, work satisfaction, better perceived work-life balance, and reduced sick leave^24^. However, potential negative effects of home working have also been reported, such as increased levels of stress and social isolation^25^. Recent reviews of the pre-pandemic literature confirmed a mixed evidence-base and highlighted existing limitations including a lack of multidimensional approaches, whereby studies consider multiple aspects of physical and psychosocial health, and sparsity of longitudinal studies ^25–27^. Nevertheless, it is unclear to what extent past evidence is translatable to home working experiences during the COVID-19 pandemic, when home working was rapidly enforced for many, and occurred in combination with other public health mitigation measures (e.g., social distancing and school closures).

Several longitudinal studies have investigated changes in workers’ mental health and wellbeing from before to during the first UK lockdown. Pelly and colleagues ^28^ identified a general pattern of improved wellbeing in workers during the first full UK lockdown (May-July 2020), including reduced levels of exhaustion and negative emotions about work. Studies have primarily suggested positive effects of working from home ^5–7^, with a few exceptions ^8^, for example, Giovanis and Ozdamar ^9^ found home working during the pandemic was associated with poorer mental health compared to those working at employer’s premises, but only for full home working, not partial home working. More recent studies have suggested that impacts of home working on mental and social wellbeing during the pandemic have been especially strong among women and mothers ^29,30^ and keyworkers ^31^. Further studies have also shown that effects may have changed over time as the pandemic has developed, though evidence for this has been mixed with one study finding the highest odds of common mental disorders among those who had consistently worked from home throughout the pandemic ^32^. Additionally, it has been reported that the initial negative impact of switching to remote working can lessen over time ^33^.

Our analyses add to this literature using data from seven ongoing UK population studies, with rich pre-pandemic information and multiple waves of data collection throughout the pandemic. Our pooled analyses have been extensively developed during the pandemic^20,34–36^ and offer considerable statistical power to examine whether associations with home working differed over time or between population sub-groups. Examining multiple indicators of mental health and social wellbeing, we provide robust evidence on the impacts of pandemic home working on specific aspects of social and mental wellbeing. Our findings confirm that associations can differ between full and partial modes of home working, between population sub-groups, and over time, showing more negative impacts at later stages of the pandemic.

Alongside the above-mentioned strengths, we note limitations. Firstly, despite adjustment for confounders, as with most observational studies, unobserved confounding could still have affected our estimates. Pre-pandemic home working was unobserved in most studies, and therefore, propensities for home working were imputed using the 2019-2020 Annual Population Survey, in combination with SOC, age group, and sex. However, sensitivity analyses conducted with data from USOC (which did collect information on pre-pandemic home working) produced similar results to those produced using imputed home working propensities. Secondly, despite adjustment for pre-pandemic wellbeing, it is possible that changes in wellbeing after measurement or during the pandemic influenced the likelihood of home working, so findings may represent reverse causality. Furthermore, while we applied study-specific weights to account for sampling design and differential non-response, residual selection bias may remain. Finally, time-points inconsistencies across sub-group analyses might indicate spurious associations and only indicate further research directions.

With many workers in the UK now maintaining home working arrangements in some form, even as COVID-19 control measures have eased, this study’s findings from the summer 2020 period, when restrictions had eased (an approximation of current conditions), can be particularly informing for working from home going forward. We found no overall association with poor social and mental wellbeing during this period, indicating that home working arrangements might continue without detrimental impacts to population mental health. As home working arrangements continue, further monitoring of mental health is essential, especially looking at sex, education, working time and age, to confirm whether these findings persist and whether some form of targeted support to protect their wellbeing may be needed.

## Conclusions

Home working became more prevalent at the start of the pandemic, but as many continue in these working patterns it is important to understand potential public health impacts. Our findings suggest that potential adverse impacts of increased home working on social and mental wellbeing are limited. We only found evidence of home working being associated with increased risk for loneliness and psychological distress during a period when lockdown measures had been re-introduced, during the winter of 2020-21. There was no overall association with such outcomes in the preceding period when restrictions had been eased. Although, there was some indication during the period of eased restrictions that partial home working may have been associated with increased risk for poor outcomes in certain population sub-groups (males, full-time workers, those with lower education, and aged 50+). Continued investigation and monitoring is advised, especially for these groups, to ensure that home working arrangements do not lead to widening inequalities in social and mental wellbeing.

## Data Availability

All datasets included in this analysis have established data sharing processes, and for most included studies the anonymised datasets with corresponding documentation can be downloaded for use by researchers from the UK Data Service. We have detailed the processes for each dataset in Supplementary File 1 and in our published protocol.

https://ukdataservice.ac.uk

## Supplementary files

Supplementary file 1. Study Description

Supplementary file 2. Data availability by time-point

Supplementary file 3. Variables description across the seven studies

Supplementary file 4. Pre-pandemic home working probabilities by occupation, age-group, and sex

Supplementary file 5. Descriptive statistics (exposure and covariates)

Supplementary file 6. Outcome descriptive by exposure

Supplementary file 7. Pooled analyses for the main results (Forest plots)

Supplementary file 8. Sub-group analyses for stratified results

Supplementary file 9. Sensitivity analyses of self-reported vs. imputed pre-pandemic home working (USOC)

Supplementary file 10. Main estimates (excluding BiB and GS)

