## Supplementary file 2 for "Home working and its association with social and mental wellbeing at different stages of the COVID-19 pandemic: Evidence from seven UK longitudinal population surveys"

**Supplementary file 2. Data availability by time-point**

|  |  | Pre-pandemic wave | 2020 | | | | | | | | | 2021 | | |
| --- | --- | --- | --- | --- | --- | --- | --- | --- | --- | --- | --- | --- | --- | --- |
|  |  |  | Apr | May | June | Jul | Aug | Sept | Oct | Nov | Dec | Jan | Feb | March |
| **CLS** | **NCDS** | 2018-2019 | Survey 1 | | | Survey 2 | | | | Survey 3 | | | | |
|  | **NS** | 2015 | Survey 1 | | | Survey 2 | | | | Survey 3 | | | | |
|  | **BCS** | 2016 | Survey 1 | | | Survey 2 | | | | Survey 3 | | | | |
| **Usoc** | | 2018-2019 | Survey  1 | Survey  2 | Survey  3 | Survey  4 |  | Survey 5 |  | Survey 6 |  | Survey 7 |  | Survey 8 |
| **ELSA** | | 2018-2019 |  |  | Survey 1 | |  |  |  | Survey 2 | |  |  |  |
| **BiB** | |  | Survey 1 | | |  |  |  | Survey 2 | |  |  |  |  |
| **GS** | | 2006-2011; 2016-2019 | Survey  1 | Survey  2 |  |  |  |  |  |  |  |  | Survey 3 |  |
| **Selected time points** | | Baseline | (T1) | | | (T2) | | | | (T3) | | | | |
