## Supplementary file 3 for "Home working and its association with social and mental wellbeing at different stages of the COVID-19 pandemic: Evidence from seven UK longitudinal population surveys"

**Supplementary file 3. Variables description across the seven studies**

1. **Outcome variables**

| Full variable name | **NS** | **BCS** | **NCDS** | **USOC** | **ELSA** | **GS** | **BIB** |
| --- | --- | --- | --- | --- | --- | --- | --- |
| Psychological Distress | GHQ-12 Scores between 0-12; cut off **4+** | Malaise Inventory (9-item version) Scores between 0-9; cut off **4+** | Malaise Inventory (9-item version) Scores between 0-9; cut off **4+** | GHQ-12 Scores between 0-12; cut off **4+** | 8-item CES-D scale; cut off **4+** | Patient Health Questionnaire (PHQ-9), Scores between 0-27 or **10+** cut off | Patient Health Questionnaire (PHQ-8), Scores between 0-27 or 10+ cut off |
| Low Life Satisfaction | Overall, how satisfied are you with your life nowadays, where 0 means 'not at all' and 10 means 'completely' - binary 0-6 | Overall, how satisfied are you with your life nowadays, where 0 means 'not at all' and 10 means 'completely' - binary 0-6 | Overall, how satisfied are you with your life nowadays, where 0 means 'not at all' and 10 means 'completely' - binary 0-6 | (2018-2019) "On a scale of 1 to 7 where 1 = 'Completely Dissatisfied' and 7 = 'Completely Satisfied', please tell me the number which you feel best describes how dissatisfied or satisfied you are with your life overall" 1=completely dissatisfied 2=Mostly disatisfied 3=Somewhat dissatisfied 4=Neither satisfied nor dissatisfied 5=Somewhat satisfied 6=Mostly satisfied 7=Completely satisfied  Recode: 1\|2\|3\|4\|5 = 1 6\|7=0 | “On a scale of 0 to 10, where 0 is “not at all” and 10 is “very”, how satisfied are you with your life nowadays?” binary 0-6 | "On a scale of 0 (not at all) to 10 (extremely), how satisfied are you with your life nowadays?" 0-6 = low | NA |
| Often Lonely | "How often do you feel lonely?" - Hardly ever / Some of the time / **Often** | "How often do you feel lonely?" - Hardly ever / Some of the time / **Often** | "How often do you feel lonely?" - Hardly ever / Some of the time / **Often** | How often do you feel lonely? 1=Hardly ever or never 2=Some of the time 3=Often  Recode: 1\|2=0 3=1 | "How often do you feel lonely?" - Hardly ever / Some of the time / **Often** | "How often have you felt lonely during the past week?" - None, or almost none of the time / Some of the time/ Most of the time / All, or almost all of the time | How often have you felt lonely during the past week? 0=None, or almost none of the time 1=Some of the time 2=Most of the time 3=All, or alsmost all of the time |
| Less than daily social contact | "In the last 7 days, how often did you talk to family or friends you do not live with via phone or video calls?" / "In the last 7 days, how often did you meet up in person with any of your family or friends you do not live with?" - Less than daily | "In the last 7 days, how often did you talk to family or friends you do not live with via phone or video calls?" / "In the last 7 days, how often did you meet up in person with any of your family or friends you do not live with?" - **Less than daily** | "In the last 7 days, how often did you talk to family or friends you do not live with via phone or video calls?" / "In the last 7 days, how often did you meet up in person with any of your family or friends you do not live with?" - **Less than daily** | Q1: In the last 4 weeks, how often have you met in person with friends and family who do not live with you?  1= less than daily 0=daily | Q1 "In the past month, how often have you done the following with any of your immediate family (parents, children, grandchildren and brothers and sisters), not counting any who live with you?" Q2. "In the past month, how often have you done the following with other relatives and/or friends?" a. Speak on the phone b. Video-Calling  1.Daily 2. 3 to 6 times a week 3. Once or twice a week 4. Less than once a week or never | "Now that COVID-19 measures are in place, how regularly do you do these activities now (W1)""How regularly do you do these activities now? (W2/3)" - Meet with family members face-to-face - Meet with friends face-to-face - Call family members - Call friends - Video call with family members (e.g., Skype, Facetime) - Video call with friends (e.g., Skype, Facetime) - Never / Rarely /Less than once a week / 1-2 days a week / 3-4 days a week / Every day/almost every day | NA |
| Fair/Poor self-rated health | "In general would you say your health is …" Excellent / Very Good / Good / Fair / Poor | "In general would you say your health is …" Excellent / Very Good / Good / **Fair / Poor** | "In general would you say your health is …" Excellent / Very Good / Good / **Fair / Poor** | In general, would you say your health is: 1. Excellent 2. Very good 3. Good 4. Fair 5. Poor  Recode: 1\|2\|3=0 4\|5=1 | “In the past month would you say your health was…” Excellent / Very good / Good / **Fair / Poor** | "In general, would you say your health is" Excellent / Very good / Good / Fair / Poor | "In general, would you say your health is" Excellent / Very good / Good / Fair / Poor |

2. **Exposure variable**

|  | **NS** | **BCS** | **NCDS** | **USOC** | **ELSA** | **GS** | **BIB** |
| --- | --- | --- | --- | --- | --- | --- | --- |
| T 1 | 0=Work from employers premises or other; 1=Work from home | 0=Work from employers premises or other; 1=Work from home | 0=Work from employers premises or other; 1=Work from home | "During the last four weeks how often did you work at home?" 1=always 2=often 3=sometimes 4=never | 1. All of my working hours are from home 2. Some of my working hours are from home 3. None of my working hours are from home | 1 = Work from own home; 2 = Work outside of the home * no partial | 0=Work from employers premises or other; 1=Work from home (no partial) |
| T 2 | 0=Work from employers premises or other; 1=Work from home; 2=Partially work from home | 0=Work from employers premises or other; 1=Work from home; 2=Partially work from home | 0=Work from employers premises or other; 1=Work from home; 2=Partially work from home | "During the last four weeks how often did you work at home?" 1=always 2=often 3=sometimes 4=never | NA | 1 = Work from own home; 2 = Work outside of the home; 3 = Partially working from home | 0=Work from employers premises or other; 1=Work from home (no partial) |
| T 3 | 0=Work from employers premises or other; 1=Work from home; 2=Partially work from home | 0=Work from employers premises or other; 1=Work from home; 2=Partially work from home | 0=Work from employers premises or other; 1=Work from home; 2=Partially work from home | "During the last four weeks how often did you work at home?" 1=always 2=often 3=sometimes 4=never | 1. All of my working hours are from home 2. Some of my working hours are from home 3. None of my working hours are from home | 1 = Work from own home; 2 = Work outside of the home; 3 = Partially working from home | NA |

3. Covariates

|  | **NS** | **BCS** | **NCDS** | **USOC** | **ELSA** | **GS** | **BIB** |
| --- | --- | --- | --- | --- | --- | --- | --- |
| Age | NA | NA | NA |  | 52+ | 27 - 100 | 16 - 57 |
| Sex | 1=Male; 2=Female | 1=Male; 2=Female | 1=Male; 2=Female | 1=Male; 2=Female | 1=Male; 2=Female | 1=Male; 2=Female | All female |
| Housing tenure | 0=Other; 1=Own/Mortgage | 0=Other; 1=Own/Mortgage | 0=Other; 1=Own/Mortgage | 0=Other; 1=Own/Mortgage | 0=Other; 1=Own/Mortgage | 0=Other; 1=Own/Mortgage | 0=Other; 1=Own/Mortgage |
| Ethnicity | 0=Non-white; 1=White | NA | NA | 1=White; 0= Non-white | 0=White; 1= Non-white | 0=White; 1= Non-white | 0=White; 1= Non-white |
| Degree level education | 0=No degree; 1=Degree | 0=No degree; 1=Degree | 0=No degree; 1=Degree | 0=no degree; 1=degree | 0=No degree; 1=Degree | 0=No degree; 1=Degree | 0=No degree; 1=Degree |
| Concurrent Household composition | 1=partner&children; 2=partner no children; 3=lone parent; 4=no partner no children; 5=alone | 1=partner&children; 2=partner no children; 3=lone parent; 4=no partner no children; 5=alone | 1=partner&children; 2=partner no children; 3=lone parent; 4=no partner no children; 5=alone | 0= no partner no children 1=no partner&children; 2=partner no children; 3=partner & children 4=alone | 1=partner&children; 2=partner no children; 3=lone parent; 4=other; 5=alone | 1=partner&children; 2=partner no children; 3=lone parent; 4=no partner no children; 5=alone | 1=partner&children; 2=partner no children; 3=lone parent; 4=no partner no children |
| Concurrent Overcrowding | number of people in household / number of rooms | number of people in household / number of rooms | number of people in household / number of rooms | number of people in household / number of rooms | number of people in household / number of rooms | number of people in household / number of rooms | number of people in household / number of rooms |
| Pre-pandemic working hours | Continuous scale (0-168) | Continuous scale (0-168) | Continuous scale (0-168) | Continuous scale (0.1 - 97.9) | Continuous scale (0-168) | For how long do YOU USUALLY Work in paid employment EACH WEEK? Hours (0: 0, 1: 1-4,2: 5-9, 3: 10-14, 4: 15-19, 5: 20-24, 6: 25-29, 7: 30-34, 8: 35-39, 9: 40-44, 10: 45-49, 11: 50-54, 12: 55-59, 13: 60+) | continuous (0-99) |
| Pre-pandemic Social Class (NSSEC 3 level) | 1= 1.1-2 (Managers); 2 = 3-4 (Intermediate); 3 = 5-9 (Lower/technical) | 1= 1.1-2 (Managers); 2 = 3-4 (Intermediate); 3 = 5-9 (Lower/technical) | 1= 1.1-2 (Managers); 2 = 3-4 (Intermediate); 3 = 5-9 (Lower/technical) | 1=management and professional 2=intermediate 3=routine | 1= 1.1-2 (Managers); 2 = 3-4 (Intermediate); 3 = 5-9 (Lower/technical); | 1=Managers, directors, senior officials; 2=Associate professional and technical occupations; 3=Administrative and secretarial occupations; 4=Skilled trades occupations; 5=Sales and customer service occupations; 6=Process, plant and machine operatives; 7=Elementary (unskilled) occupations; 8=Never worked | 1=Modern professional; 2=Clerical and intermediate; 3=Senior managers/administrators; 4 =Technical and craft; 5=Semi-routine manual; 6 =Routine manual and service; 7=Middle/junior managers; 8=Traditional professional; 9=Other |
| Pre-pandemic home working (more or less likely to work from home) | <https://www.ons.gov.uk/employmentandlabourmarket/peopleinwork/labourproductivity/datasets/homeworkingintheukworkfromhomestatus> | (2012) Yes, Work from home, No fixed place of work | (2008) Yes, Work from home, No fixed place of work | 0=never 1=partial wfh 2= always wfh |  | From COVID Wave 2: During January and February how often did you work at home? (4 - Always; 3 - Often; 2 - Sometimes; 1 - Never; 97 - Not applicable) | Pre-pandemic: How do you usually travel to work (option: I mainly work from home). Pandemic: Do you currently work from home? |
| Pre-pandemic Occupation (1-digit SOC) 9 level | 1)Managers, Directors And Senior Officials 2)Professional Occupations 3)Associate Professional And Technical Occupations 4)Administrative And Secretarial Occupations 5)Skilled Trades Occupations 6)Caring, Leisure And Other Service Occupations 7)Sales And Customer Service Occupations 8)Process, Plant And Machine Operatives 9)Elementary Occupations | 1)Managers, Directors And Senior Officials 2)Professional Occupations 3)Associate Professional And Technical Occupations 4)Administrative And Secretarial Occupations 5)Skilled Trades Occupations 6)Caring, Leisure And Other Service Occupations 7)Sales And Customer Service Occupations 8)Process, Plant And Machine Operatives 9)Elementary Occupations | 1)Managers, Directors And Senior Officials 2)Professional Occupations 3)Associate Professional And Technical Occupations 4)Administrative And Secretarial Occupations 5)Skilled Trades Occupations 6)Caring, Leisure And Other Service Occupations 7)Sales And Customer Service Occupations 8)Process, Plant And Machine Operatives 9)Elementary Occupations | Three digit SOC 2010 | 1)Managers, Directors And Senior Officials 2)Professional Occupations 3)Associate Professional And Technical Occupations 4)Administrative And Secretarial Occupations 5)Skilled Trades Occupations 6)Caring, Leisure And Other Service Occupations 7)Sales And Customer Service Occupations 8)Process, Plant And Machine Operatives 9)Elementary Occupations | NA | Not available. |
| Concurrent Key worker status | 1=Yes; 2=No | 1=Yes; 2=No | 1=Yes; 2=No | 1=yes; 2=no | 1=Yes; 2=No | 1=Yes; 2=No | 1=Yes; 2=No |
| Psychological distress | (2015) GHQ-12 **>=4** | (2016) Malaise Inventory >=4 | (2008) Malaise inventory **>=4** | (2018-2019) GHQ-12 >=4 | (2018/19)  CES-D score >=4 | (2006 - 2011) GHQ-28 > = 24 | Patient Health Questionnaire (PHQ-8),Scores between 0-15 or 10+ cut off |
| Low life satisfaction | (2015) W8OSATIS Overall life satisfaction: **Neither satisfied dissatisfied or Very dissatisfied** | (2016) B10LIFST1 Life satisfaction (0-10 scale): **0-6** | (2008) N8LIFET1 Life satisfaction (0-10 scale): **0-6** | (2018-2019) "On a scale of 1 to 7 where 1 = 'Completely Dissatisfied' and 7 = 'Completely Satisfied', please tell me the number which you feel best describes how dissatisfied or satisfied you are with your life overall" 1=completely dissatisfied 2=Mostly disatisfied 3=Somewhat dissatisfied 4=Neither satisfied nor dissatisfied 5=Somewhat satisfied 6=Mostly satisfied 7=Completely satisfied  Recode: 1\|2\|3\|4\|5 = 1 6\|7=0 | (2018/19)  Life satisfaction (0-10 scale): 0-6 | (2020, retrospective) Thinking back to just before the COVID-19 measures were introduced (i.e., January 2020), how satisfied were you with your life then? 0 being not at all, 10 being extremely | NA |
| Self-rated health | (2015) W8GENA "Describe your health generally": **Fair or Poor** | (2016) B10HLTHGN "Describe your health generally": **Fair or Poor** | (2013) "Describe your health generally": **Fair or Poor** | (2018-2019) In general, would you say your health is: 1=Excellent 2=Very good 3=Good 4=Fair 5=Poor  Recode: 1\|2\|3 = 0 4\|5=1 | (2018/19) Fair/poor | NA | "How would you describe your health generally". Five-point ordinal scale from 1 (Excellent) to 5 (Poor) |
| Loneliness | (2015) W8TALK "People around willing to listen to problems": **Not at all or a little** | (2016) B10Q1I "Feels close to others": **Rarely or Never** | (2008) N8LISTEN "People around to listen to problems": Not at all or little | (2018-2019) How often do you feel lonely? 1=Hardly ever or never 2=Some of the time 3=Often  Recode: 1\|2=0 3=1 | (2018/19) How often do you feel lonely? - Hardly ever / Some of the time / Often | (2006 - 2011) Do you often feel lonely? Yes/No | NA |
| Less than daily social contact | (2015) W8FAMMT W8FREMT "How often meets family/friends": **Less than weekly** | (2016) B10FAMMT B10FREMT "How often meets family/friends": **Less than three or more times a week** | (2008) N8VISITA N8VISITB "How often sees friends": **Less than 3-6 times each fortnight** | NA | (2018/19) "How often do you meet up with children/ other family members/ friends on average?" **Less than three times a week** | (2020, retrospective) "Just before COVID-19 measures were introduced (i.e., January 2020), how regularly did you:" - Meet with family members face-to-face - Meet with friends face-to-face - Call family members - Call friends - Video call with family members (e.g., Skype, Facetime) - Video call with friends (e.g., Skype, Facetime) - Never / Rarely /Less than once a week / 1-2 days a week / 3-4 days a week / Every day/almost every day | NA |
| Chronic health disabilities | (2015) W8LOIL "Longstanding illness": **Yes** | (2016) B10LOIL "Long-lasting physical or mental illnesses": **Yes** | (2013) N9LOIL "Long-lasting physical or mental illnesses": **Yes** | (2018-2019) "Do you have any long-standing physical or mental impairment, illness or disability? By 'long-standing' I mean anything that has troubled you over a period of at least 12 months or that is likely to trouble you over a period of at least 12 months" 1=yes 0=no | (2018/19) "Long-standing limiting illness" **Yes** | NA | "Do you have a long standing illness, disability or infirmity" yes/no/does not wish to answer |
