## Supplementary file 4 for "Home working and its association with social and mental wellbeing at different stages of the COVID-19 pandemic: Evidence from seven UK longitudinal population surveys"

**Supplementary file 4. Pre-pandemic home working probabilities by occupation, age-group and sex**

We control for pre-pandemic home working, but this information is not contained in all the datasets used in this study. We therefore derive the propensities to work fully or partially from home prior the start of the pandemic from an external dataset, the Annual Population Survey (APS). The Office for National Statistics (ONS) has shown, using the APS for the period April 2019 to March 2020, differences across occupational classification (SOC2010, 1 digit) by home working (Office for National Statistics, no date). We replicate the analyses using SOC2010 2 digits as well as sex and age as explanatory variables and derive the propensities from such a model.

**Dataset and analyses**

Annual Population Survey (APS), April 2019 - March 2020

Principal Investigator: Office for National Statistics, Social Survey Division

Reference: Office for National Statistics, Social Survey Division. (2021). Annual Population Survey, April 2019 - March 2020. [data collection]. 6th Edition. UK Data Service. SN: 8647, <http://doi.org/10.5255/UKDA-SN-8647-6> Variables information can be found on the UK data service website.

**Variables used**

***Home working*** in main job (HOME), distinguishing those working fully from the workplace from those not working fully from the workplace (binary).

***Age*** (AGE) recoded as three age-groups 16-29, 30-49 and 50-66

***Sex*** (SEX) coded as male and female

Three digits of the ***Standard Occupational Classification***, SOC2010 (SC10MMN) recoded on a 2-digit basis (excluding the third digit).

**Model**

Binary logit regression calculating the propensities (in logit) to work fully or partially from home versus not working from home (reference category). Data are weighted using a standardized population weight variable (PWTA20). The model includes three interaction terms: between SOC2010 and sex, between SOC2010 and age group and between sex and age group. Predicted probabilities are derived from the logits.

Predicted probabilities are then merged with the different dataset used in the study. Sensitivity analyses will be performed with the datasets for which information about pre-pandemic home working has been collected to ensure the validity of the data.

**Results**

|  | Estimate | Std. Error | z value | Pr(>\|z\|) |
| --- | --- | --- | --- | --- |
| (Intercept) | -4.4588 | 0.1838 | -24.2580 | 0.0000 |
| sexmale | -0.1510 | 0.1924 | -0.7850 | 0.4325 |
| agegroup30-49 | 0.3445 | 0.2189 | 1.5739 | 0.1155 |
| agegroup50-66 | 0.0730 | 0.2389 | 0.3053 | 0.7601 |
| SOC_2_11 | 1.7205 | 0.2523 | 6.8190 | 0.0000 |
| SOC_2_12 | 0.3540 | 0.4376 | 0.8089 | 0.4186 |
| SOC_2_21 | 1.7519 | 0.2267 | 7.7286 | 0.0000 |
| SOC_2_22 | -0.4975 | 0.4542 | -1.0954 | 0.2733 |
| SOC_2_23 | -0.3875 | 0.4180 | -0.9269 | 0.3540 |
| SOC_2_24 | 1.0754 | 0.2555 | 4.2081 | 0.0000 |
| SOC_2_31 | 0.8741 | 0.3546 | 2.4649 | 0.0137 |
| SOC_2_32 | 0.9351 | 0.3881 | 2.4092 | 0.0160 |
| SOC_2_33 | -0.9635 | 1.0563 | -0.9122 | 0.3617 |
| SOC_2_34 | 1.3440 | 0.3058 | 4.3955 | 0.0000 |
| SOC_2_35 | 1.5065 | 0.2097 | 7.1832 | 0.0000 |
| SOC_2_41 | 0.8731 | 0.2345 | 3.7227 | 0.0002 |
| SOC_2_42 | 0.6701 | 0.3654 | 1.8338 | 0.0667 |
| SOC_2_51 | 3.1670 | 0.5170 | 6.1260 | 0.0000 |
| SOC_2_52 | 0.9930 | 0.4840 | 2.0515 | 0.0402 |
| SOC_2_53 | 2.1664 | 0.5549 | 3.9039 | 0.0001 |
| SOC_2_54 | -0.0916 | 0.5187 | -0.1765 | 0.8599 |
| SOC_2_61 | 1.0569 | 0.2232 | 4.7359 | 0.0000 |
| SOC_2_62 | -0.0097 | 0.4375 | -0.0221 | 0.9823 |
| SOC_2_71 | -1.0890 | 0.3445 | -3.1612 | 0.0016 |
| SOC_2_72 | -0.4733 | 0.4180 | -1.1321 | 0.2576 |
| SOC_2_81 | 0.3196 | 0.4337 | 0.7370 | 0.4611 |
| SOC_2_82 | 0.7148 | 0.7196 | 0.9933 | 0.3206 |
| SOC_2_91 | 1.9796 | 0.3279 | 6.0366 | 0.0000 |
| sexmale:agegroup30-49 | 0.1561 | 0.1034 | 1.5091 | 0.1313 |
| sexmale:agegroup50-66 | 0.3528 | 0.1073 | 3.2883 | 0.0010 |
| sexmale:SOC_2_11 | -0.1624 | 0.1943 | -0.8358 | 0.4033 |
| sexmale:SOC_2_12 | 0.0243 | 0.2299 | 0.1056 | 0.9159 |
| sexmale:SOC_2_21 | 0.0328 | 0.2047 | 0.1604 | 0.8726 |
| sexmale:SOC_2_22 | 0.0105 | 0.2897 | 0.0361 | 0.9712 |
| sexmale:SOC_2_23 | 0.2032 | 0.2482 | 0.8188 | 0.4129 |
| sexmale:SOC_2_24 | 0.1720 | 0.2014 | 0.8542 | 0.3930 |
| sexmale:SOC_2_31 | 0.0205 | 0.2849 | 0.0721 | 0.9426 |
| sexmale:SOC_2_32 | -0.0877 | 0.2971 | -0.2952 | 0.7678 |
| sexmale:SOC_2_33 | -0.2603 | 0.4965 | -0.5242 | 0.6001 |
| sexmale:SOC_2_34 | 0.0236 | 0.2714 | 0.0871 | 0.9306 |
| sexmale:SOC_2_35 | 0.2134 | 0.1931 | 1.1053 | 0.2690 |
| sexmale:SOC_2_41 | -0.3259 | 0.2129 | -1.5309 | 0.1258 |
| sexmale:SOC_2_42 | -0.0249 | 0.3851 | -0.0646 | 0.9485 |
| sexmale:SOC_2_51 | -0.5391 | 0.4811 | -1.1206 | 0.2625 |
| sexmale:SOC_2_52 | 0.2976 | 0.4610 | 0.6456 | 0.5185 |
| sexmale:SOC_2_53 | 0.2146 | 0.5467 | 0.3926 | 0.6946 |
| sexmale:SOC_2_54 | 0.4135 | 0.4275 | 0.9673 | 0.3334 |
| sexmale:SOC_2_61 | 0.1499 | 0.2405 | 0.6232 | 0.5331 |
| sexmale:SOC_2_62 | 0.1680 | 0.3242 | 0.5183 | 0.6043 |
| sexmale:SOC_2_71 | 0.9257 | 0.2909 | 3.1822 | 0.0015 |
| sexmale:SOC_2_72 | 0.8496 | 0.2934 | 2.8957 | 0.0038 |
| sexmale:SOC_2_81 | 0.1249 | 0.3431 | 0.3640 | 0.7159 |
| sexmale:SOC_2_82 | 0.0796 | 0.6643 | 0.1199 | 0.9046 |
| sexmale:SOC_2_91 | -0.1233 | 0.3177 | -0.3880 | 0.6980 |
| agegroup30-49:SOC_2_11 | 0.5367 | 0.2770 | 1.9374 | 0.0527 |
| agegroup50-66:SOC_2_11 | 1.1407 | 0.2932 | 3.8900 | 0.0001 |
| agegroup30-49:SOC_2_12 | 1.6668 | 0.4562 | 3.6541 | 0.0003 |
| agegroup50-66:SOC_2_12 | 2.0622 | 0.4693 | 4.3944 | 0.0000 |
| agegroup30-49:SOC_2_21 | 0.4045 | 0.2479 | 1.6318 | 0.1027 |
| agegroup50-66:SOC_2_21 | 0.9127 | 0.2701 | 3.3791 | 0.0007 |
| agegroup30-49:SOC_2_22 | 0.6605 | 0.4882 | 1.3531 | 0.1760 |
| agegroup50-66:SOC_2_22 | 1.6150 | 0.4954 | 3.2598 | 0.0011 |
| agegroup30-49:SOC_2_23 | 0.7518 | 0.4469 | 1.6821 | 0.0926 |
| agegroup50-66:SOC_2_23 | 1.7177 | 0.4569 | 3.7592 | 0.0002 |
| agegroup30-49:SOC_2_24 | 0.7343 | 0.2818 | 2.6056 | 0.0092 |
| agegroup50-66:SOC_2_24 | 1.5063 | 0.2986 | 5.0439 | 0.0000 |
| agegroup30-49:SOC_2_31 | 0.2873 | 0.3672 | 0.7824 | 0.4340 |
| agegroup50-66:SOC_2_31 | 1.1228 | 0.3795 | 2.9582 | 0.0031 |
| agegroup30-49:SOC_2_32 | 0.3171 | 0.4316 | 0.7348 | 0.4624 |
| agegroup50-66:SOC_2_32 | 1.3978 | 0.4324 | 3.2323 | 0.0012 |
| agegroup30-49:SOC_2_33 | 1.2159 | 1.0656 | 1.1410 | 0.2539 |
| agegroup50-66:SOC_2_33 | 2.0673 | 1.0829 | 1.9090 | 0.0563 |
| agegroup30-49:SOC_2_34 | 0.5896 | 0.3389 | 1.7398 | 0.0819 |
| agegroup50-66:SOC_2_34 | 1.4703 | 0.3716 | 3.9565 | 0.0001 |
| agegroup30-49:SOC_2_35 | 0.7017 | 0.2401 | 2.9230 | 0.0035 |
| agegroup50-66:SOC_2_35 | 1.2184 | 0.2607 | 4.6737 | 0.0000 |
| agegroup30-49:SOC_2_41 | 0.6119 | 0.2680 | 2.2829 | 0.0224 |
| agegroup50-66:SOC_2_41 | 0.9891 | 0.2859 | 3.4589 | 0.0005 |
| agegroup30-49:SOC_2_42 | 0.8268 | 0.4049 | 2.0418 | 0.0412 |
| agegroup50-66:SOC_2_42 | 1.1820 | 0.4130 | 2.8623 | 0.0042 |
| agegroup30-49:SOC_2_51 | -0.4017 | 0.3981 | -1.0090 | 0.3130 |
| agegroup50-66:SOC_2_51 | -0.3452 | 0.4255 | -0.8114 | 0.4171 |
| agegroup30-49:SOC_2_52 | 0.4991 | 0.2932 | 1.7022 | 0.0887 |
| agegroup50-66:SOC_2_52 | 0.6741 | 0.3144 | 2.1440 | 0.0320 |
| agegroup30-49:SOC_2_53 | -0.2320 | 0.2909 | -0.7973 | 0.4253 |
| agegroup50-66:SOC_2_53 | -0.2960 | 0.3228 | -0.9170 | 0.3592 |
| agegroup30-49:SOC_2_54 | 0.1145 | 0.4935 | 0.2321 | 0.8165 |
| agegroup50-66:SOC_2_54 | 0.0336 | 0.5524 | 0.0607 | 0.9516 |
| agegroup30-49:SOC_2_61 | -0.2900 | 0.2682 | -1.0814 | 0.2795 |
| agegroup50-66:SOC_2_61 | 0.3796 | 0.2829 | 1.3420 | 0.1796 |
| agegroup30-49:SOC_2_62 | 0.6013 | 0.4999 | 1.2030 | 0.2290 |
| agegroup50-66:SOC_2_62 | 1.7088 | 0.4891 | 3.4937 | 0.0005 |
| agegroup30-49:SOC_2_71 | 0.6683 | 0.3812 | 1.7532 | 0.0796 |
| agegroup50-66:SOC_2_71 | 1.4635 | 0.3865 | 3.7860 | 0.0002 |
| agegroup30-49:SOC_2_72 | 1.0958 | 0.4392 | 2.4949 | 0.0126 |
| agegroup50-66:SOC_2_72 | 1.5653 | 0.4652 | 3.3652 | 0.0008 |
| agegroup30-49:SOC_2_81 | 0.2718 | 0.4177 | 0.6507 | 0.5152 |
| agegroup50-66:SOC_2_81 | 0.4576 | 0.4377 | 1.0454 | 0.2958 |
| agegroup30-49:SOC_2_82 | -0.6192 | 0.4583 | -1.3511 | 0.1767 |
| agegroup50-66:SOC_2_82 | -0.3494 | 0.4598 | -0.7597 | 0.4474 |
| agegroup30-49:SOC_2_91 | -0.5813 | 0.3367 | -1.7266 | 0.0842 |
| agegroup50-66:SOC_2_91 | -0.7646 | 0.3942 | -1.9394 | 0.0524 |

Predicted probability were derived from these estimates.

ELSA does not have the SOC2010 classification but only the SOC2000 and the Annual Population Survey only has the SOC2010 classification but not the SOC2000. To tackle this issue, we have used the probabilities to be classified in SOC2010 based on SOC2000 and multiplied these probabilities with the predicted probabilities. SOC2010/SOC2000 probabilities come from:

Elias P., Birch M., [SOC2010 The revision of the Standard Occupational Classification 2000](https://www.google.com/url?sa=t&rct=j&q=&esrc=s&source=web&cd=&ved=2ahUKEwi0iqCnjOH0AhUQi1wKHXoFB7wQFnoECAoQAQ&url=https%3A%2F%2Fwww.ons.gov.uk%2Ffile%3Furi%3D%2Fmethodology%2Fclassificationsandstandards%2Fstandardoccupationalclassificationsoc%2Fsoc2010%2Fsoc2010volume1structureanddescriptionsofunitgroups%2Frevisionofsoc2000paperpeliasfinaltcm77181318.pdf&usg=AOvVaw3th3PJ7owBs4zgkKuoTK2f).

USOC has a mixture of SOC2000 and SOC2010 classifications, available dependent upon when participants last changed employment. To tackle this issue, we included a dummy variable in any analyses that included pre-pandemic home working (derived from the APS) to indicate whether SOC2000 or SOC2010 was used.
