## Supplementary file 5 for "Home working and its association with social and mental wellbeing at different stages of the COVID-19 pandemic: Evidence from seven UK longitudinal population surveys"

**Supplementary file 5. Descriptive statistics (exposure and covariates)**

|  | **NS** | **BCS** | **NCDS** | **ELSA** | **USOC** | **GS** | **BiB** |
| --- | --- | --- | --- | --- | --- | --- | --- |
| **Wave 1** | May-20 | May-20 | May-20 | June/July 20 weighted | May 20 (weighted) |  | Apr - Jun 2020 |
| Proportion wfh | 65.5 (651) | 56.5 (1114) | 42.2 (573) | 32.89 (224) | 59.3 (2167.7) | 59.8 (1014) | 48.7 (180) |
| Working hours wfh - Mean (SD) | 35.4 (10.9) | 35.5 (11.4) | 32.3 (11.3) | 34.25 (15.04) | 32.9 (14.1) | / | - |
| Proportion NOT wfh | 34.5 (292) | 43.5 (759) | 57.8 (596) | 54.16 (339) | 40.7 (1490) | 34.2 (579) | 51.4 (190) |
| Working hours NOT wfh - Mean (SD) | 34.5 (13.6) | 35.4 (14.0) | 33.6 (13.9) | 31.72 (11.43) | 30.7 (16.3) | / |  |
| ***Covariates*** |  |  |  |  |  |  |  |
| Age, mean (range) | 30 | 50 | 62 | 58.46 (52-66) | 43.6 (18-66) | 50.24 (27 - 66) | 41.3 (28 - 55) |
| Sex (Female) | 49.0 (575) | 49.8 (1083) | 51.1 (620) | 46.87 (367) | 53.6 (1961.1) | 65.5 (1110) | 100 (370) |
| Housing tenure (owns home) | 23.9 (231) | 83.3 (1629) | 85.9 (1053) | 82.66 (585) | 73.2 (2676.3) | 90.9 (1679) | 76.1 (277) |
| Ethnicity (White) | 87.3 (779) | / | / | 89.2 (624) | 92.7 (3389.4) | 99.1 (1679) | 57.5 (207) |
| Degree level education | 52.2 (553) | 44.9 (1023) | 36.8 (538) | 28.03 (214) | 51.5 (1883.1) | 58.1 (985) | 50.3 (170) |
| pre_Working hours - Mean (SD) | 38.4 (9.6) | 37.7 (10.3) | 34.0 (11.2) | 35.06 (12.39) | 35.8 (11.4) | 33.45 | 30..2 (12.5) |
| pre_Working from home | 0.08 (0.05) | 0.11 (0.07) | 0.09 (0.06) | 0.08 (0.05) | 3.1 (114.9) | 0.3 (57) | 2.0 (7) |
| Socio-economic classification (1) | 64.4 (654) | 56.2 (1144) | 41.2 (574) | 30.77 (270) | 51.5 (1882.8) | 63.5 (1077) | - |
| Socio-economic classification (2) | 20.3 (174) | 24.0 (435) | 27.3 (298) | 13.88 (121) | 17.5 (640) | 21.8 (369) | - |
| Socio-economic classification (3) | 15.4 (115) | 19.8 (294) | 31.6 (297) | 25.03 (193) | 31.0 (1134.9) | 9.9 (167) | - |
| Managers, directors and senior SOC (1) | 5.4 (67) | 14.3 (283) | 10.4 (139) | 15.08 (107) | 11.3 (414) | NA | See comment |
| Professional SOC (2) | 39.9 (385) | 27.8 (596) | 20.2 (292) | 13.63 (94) | 22.6 (826.6) | NA |  |
| Associate professional and technical SOC (3) | 23.6 (235) | 17.7 (326) | 12.1 (157) | 15.61 (106) | 19.1 (697.5) | NA |  |
| Administrative and secretarial SOC (4) | 10.4 (104) | 14.2 (266) | 17.9 (208) | 15.32 (99) | 11.4 (416.1) | NA |  |
| Skilled trades SOC (5) | 3.7 (19) | 6.0 (79) | 7.3 (80) | 10.91 (59) | 5.9 (215.8) | NA |  |
| Caring, leisure and other service SOC (6) | 7.8 (49) | 6.9 (137) | 14.1 (119) | 8.84 (58) | 9.7 (355.3) | NA |  |
| Sales and customer service SOC (7) | 4.4 (41) | 4.3 (76) | 5.8 (65) | 4.32 (33) | 7.7 (281.3) | NA |  |
| Process, plant and machine operatives SOC (8) | 2.0 (20) | 4.6 (55) | 6.0 (61) | 5.72 (43) | 3.8 (139.2) | NA |  |
| Elementary occupations SOC (9) | 2.8 (23) | 4.2 (55) | 6.2 (48) | 10.58 (60) | 8.5 (311.8) | NA |  |
| Key worker | 55.9 (504) | 57.3 (1079) | 65.1 (719) | 49.15 (321) | 58.3 (2133.0) | 53.2 (901) | 72.7 (269) |
| HH (Female, partner + children) | 13.8 (117) | 27.3 (632) | 8.6 (113) | 14.42 (102) | 15.1 (551.1) | 29.7 (503) | 71.1 (256) |
| HH (Male, partner + children) | 10.8 (66) | 30.2 (512) | 15.0 (170) | 19.55 (99) | 16.8 (613.7) | 16.3 (276) | NA |
| HH (Female, partner no children) | 21.5 (253) | 8.7 (189) | 22.2 (274) | 18.91 (174) | 20.5 (751.5) | 21.2 (359) | 0.3 (1) |
| HH (Male, partner no children) | 23.5 (169) | 7.8 (125) | 21.2 (251) | 24.97 (142) | 18.2 (664.8) | 12.6 (213) |  |
| HH (lone parents) | 1.9 (24) | 7.3 (132) | 4.4 (53) | 5.86 (40) | 3.7 (135.9) | 6.0 (101) | 28.6 (103) |
| HH (others, e.g., housemates) | 18.7 (189) | 5.2 (71) | 6.0 (59) | 2.56 (20) | 14.8 (541.5) | 2.1 (36) | 0 |
| HH (alone) | 9.8 (125) | 13.4 (212) | 22.6 (249) | 13.73 (82) | 10.9 (399.2) | 12.2 (207) | 0 |
| Overcrowding - Mean (SD) | 0.7 (0.4) | 0.6 (0.6) | 0.5 (0.3) | 0.53 (0.28) | 0.6 (0.3) | .46 (.22) | 1.6 (0.5) |
| pre_Chronic health disability | 16.6 (173) | 31.9 (603) | 27.1 (321) | 15.06 (91) | 25.8 (943) |  | 25.1 (93) |
| pre_Psychological Distress | 22.7 (238) | 14.1 (240) | 10.5 (118) | 8.46 (43) | 17.8 (652.9) | 9.4 (160) | 10 (37) |
| pre_Low Life Satisfaction | 20.2 (198) | 9.8 (162) | 21.4 (236) | 26.53 (142) | 23.1 (845.8) | 13.5 (229) | - |
| pre_Fair/Poor Self-rated health | 6.3 (54) | 13.8 (209) | 11.8 (119) | 18.41 (79) | 13.3 (486.8) | - | 19.5 (72) |
| pre_Often Lonely | 7.1 (88) | 6.2 (101) | 8.0 (76) | 3.76 (21) | 7.3 (265.5) | 9.2 (156) | - |
| pre_Less than daily Social Contact | 71.4 (717) | 69.4 (1343) | 65.5 (794) | 34.26 (223) |  | 31.3 (530) | - |
| **Total N** | 943 | 1873 | 1169 | 659 | 3658 | 1695 | 370 |
| **Wave 2** | Sep-20 | Sep-20 | Sep-20 | NA | Sept 20 (weighted) |  | Oct 2020 - Jan 2021 |
| Proportion fully wfh | 27.8 (667) | 25.7 (748) | 18.5 (355) |  | 23.4 (918.1) | 41.3 (496) | 64.7 (88) |
| Working hours fully wfh - Mean (SD) | 38.4 (7.3) | 39.2 (9.4) | 34.1 (11.4) |  | 35.3 (13.0) | / | - |
| Proportion partially wfh | 16.8 (320) | 15.3 (419) | 10.3 (208) |  | 22.0 (861.3) | 18.7 (224) | - |
| Working hours partially wfh - Mean (SD) | 40.5 (10.5) | 39.2 (9.4) | 35.1 (11.7) |  | 36.1 (12.7) | / | - |
| Proportion not wfh | 55.4 (1001) | 59.0 (1447) | 71.2 (1166) |  | 54.6 (2138.3) | 29.3 (351) | 35.3 (48) |
| Working hours not wfh - Mean (SD) | 38.0 (11.7) | 37.6 (11.4) | 33.4 (12.0) |  | 33.7 (14.5) | / | - |
| ***Covariates*** |  |  |  |  |  |  |  |
| Age/Age range | 30 | 50 | 62 |  | 43.3 (18-66) | 50.9 (27 - 66) | 42 (28 - 54) |
| Sex (Female) | 48.1 (1213) | 48.9 (1470) | 51.0 (930) |  | 53.7 (2101.9) | 66.3 (796) | 100 (136) |
| Housing tenure (owns home) | 22.1 (450) | 80.0 (2214) | 84.8 (1520) |  | 69.6 (2725.7) | 90.8 (1082) | 80.0 (108) |
| Ethnicity (White) | 88.3 (1529) | / | / |  | 92.8 (3634.4) | 98.9 (1187) | 67.2 (88) |
| Degree level education | 46.4 (1047) | 42.7 (1297) | 31.6 (677) |  | 47.9 (1878) | 59.2 (710) | 58.6 (75) |
| pre_Working hours - Mean (SD) | 38.6 (9.8) | 38.0 (10.4) | 34.1 (11.6) |  | 35.7 (11.5) | 33.5 | 30.4 (11.1) |
| pre_Working from home | 0.07 (0.05) | 0.10 (0.07) | 0.09 (0.06) |  | 2.8 (111.0) | 4.8 (57) | 2.3 (3) |
| Socio-economic classification (1) | 56.0 (1204) | 50.5 (1442) | 37.4 (730) |  | 48.2 (1887.2) | 63.0 (756) | - |
| Socio-economic classification (2) | 24.8 (449) | 23.0 (615) | 23.4 (434) |  | 16.9 (661.2) | 22.2 (266) | - |
| Socio-economic classification (3) | 19.1 (335) | 26.5 (557) | 39.2 (565) |  | 35.0 (1369.3) | 10.3 (123) | - |
| Managers, directors and senior SOC (1) | 10.7 (149) | 16.1 (398) | 11.0 (188) |  | 11.0 (432.3) | NA | See comment |
| Professional SOC (2) | 27.1 (651) | 21.7 (691) | 15.1 (336) |  | 19.6 (767) | NA |  |
| Associate professional and technical SOC (3) | 22.4 (468) | 15.7 (430) | 12.5 (221) |  | 18.1 (709.4) | NA |  |
| Administrative and secretarial SOC (4) | 11.2 (237) | 13.4 (364) | 15.1 (291) |  | 11.2 (437.4) | NA |  |
| Skilled trades SOC (5) | 6.5 (75) | 7.4 (150) | 9.2 (148) |  | 8.1 (317.4) | NA |  |
| Caring, leisure and other service SOC (6) | 8.1 (163) | 9.3 (236) | 13.8 (202) |  | 9.4 (369.5) | NA |  |
| Sales and customer service SOC (7) | 6.5 (126) | 4.4 (116) | 6.2 (113) |  | 8.6 (337) | NA |  |
| Process, plant and machine operatives SOC (8) | 3.2 (45) | 7.3 (123) | 8.5 (110) |  | 4.3 (167.2) | NA |  |
| Elementary occupations SOC (9) | 4.3 (74) | 4.7 (106) | 8.5 (120) |  | 9.7 (380.5) | NA |  |
| Key worker | 50.5 (984) | 52.6 (1367) | 54.3 (922) |  | 50.1 (1962.8) | 52.2 (626) | - |
| HH (Female, partner + children) | 13.5 (285) | 24.3 (792) | 7.6 (152) |  | 14.7 (540.6) | 28.8 (346) | 30.2 (41) |
| HH (Male, partner + children) | 13.1 (166) | 28.6 (686) | 10.8 (189) |  | 16.5 (605.0) | 15.7 (188) | NA |
| HH (Female, partner no children) | 16.7 (448) | 9.0 (269) | 22.2 (424) |  | 20.5 (752.4) | 22.4 (269) | 0 |
| HH (Male, partner no children) | 17.7 (298) | 9.9 (211) | 2.5 (378) |  | 18.2 (667.3) | 12.6 (151) | NA |
| HH (lone parents) | 4.2 (79) | 8.3 (214) | 4.2 (76) |  | 3.8 (138.9) | 6.4 (77) | 30.2 (41) |
| HH (no partner no children) | 17.8 (434) | 2.9 (54) | 2.7 (37) |  | 15.7 (574.9) | 1.9 (23) | 0 |
| HH (alone) | 16.8 (278) | 17.0 (388) | 29.0 (473) |  | 10.6 (390.4) | 12.2 (146) | 0 |
| Overcrowding - Mean (SD) | 0.8 (0.4) | 0.6 (0.4) | 0.5 (0.3) |  | 0.7 (0.3) | .45 (.22) | 1.5 (0.4) |
| pre_Chronic health disability | 15.3 (353) | 30.3 (839) | 28.4 (479) |  | 27.3 (1068.5) | - | 22.1 (30) |
| pre_Psychological Distress | 21.7 (498) | 15.7 (368) | 10.3 (181) |  | 17.0 (664.4) | 9.1 (109) | 8.8 (12) |
| pre_Low Life Satisfaction | 21.8 (461) | 12.6 (279) | 21.8 (361) |  | 24.7 (968.7) | 13.3 (159) | - |
| pre_Fair/Poor Self-rated health | 6.9 (135) | 15.9 (345) | 14.7 (201) |  | 13.6 (534.6) | - | 16.9 (23 |
| pre_Often Lonely | 7.0 (191) | 6.8 (161) | 7.0 (116) |  | 7.7 (302.1) | 8.9 (107) |  |
| pre_Less than daily Social Contact | 66.0 (1147) | 69.8 (1861) | 65.7 (1147) |  | - | 31.0 (372) |  |
| **Total N** | 1988 | 2614 | 1729 |  | 3918 | 1200 | 136 |
| **Wave 3** | Feb-21 | Feb-21 | Feb-21 | Nov/Dec 20 weighted | Jan 21 (weighted) |  | NA |
| Proportion fully wfh | 37.8 (996) | 33.3 (1086) | 24.1 (515) | 23.15 (178) | 36.8 (1231.7) | 42.9 (466) |  |
| Working hours fully wfh - Mean (SD) | 39.3 (8.4) | 38.9 (9.6) | 34.4 (15.4) | 33.12 (12.22) | 36.6 (13.5) | / |  |
| Proportion partially wfh | 14.5 (369) | 16.2 (497) | 12.4 (233) | 16.54 (124) | 17.6 (589.5) | 21.3 (232) |  |
| Working hours partially wfh - Mean (SD) | 38.3 (8.7) | 39.5 (10.5) | 35.2 (11.6) | 35.21 (13.80) | 36.2 (11.1) | / |  |
| Proportion not wfh | 47.6 (936) | 50.5 (1338) | 63.4 (1065) | 60.31 (407) | 45.6 (1526.4) | 23.6 (257) |  |
| Working hours not wfh - Mean (SD) | 36.8 (13.5) | 37.2 (11.8) | 32.5 (11.5) | 33.23 (11.03) | 34.0 (15.2) | / |  |
| ***Covariates*** |  |  |  |  |  |  |  |
| Age/Age range | 31 | 51 | 63 | 58.61 (52-66) | 44.0 (18-66) | 51.1 (28 - 66) |  |
| Sex (Female) | 49.4 (1355) | 49.0 (1646) | 49.0 (976) | 50.42 (409) | 50.6 (1693.3) | 65.2 (709) |  |
| Housing tenure (owns home) | 20.6 (509) | 81.3 (2486) | 86.5 (1607) | 82.57 (629) | 72.1 (2413.9) | 90.9 (988) |  |
| Ethnicity (White) | 70.3 (1746) | / | / | 91.57 (674) | 92.6 (3101.5) | 99.1 (1077) |  |
| Degree level education | 48.0 (1248) | 45.8 (1485) | 33.5 (716) | 26.67 (227) | 49.0 (1641.7) | 57.6 (626) |  |
| pre_Working hours - Mean (SD) | 38.5 (9.4) | 37.3 (10.4) | 33.6 (11.3) | 34.08 (11.58) | 36.5 (11.1) | 33.8 |  |
| pre_Working from home | 0.07 (0.05) | 0.10 (0.07) | 0.09 (0.06) | 0.08 (0.05) | 3.1 (103.5) | 4.3 (47) |  |
| Socio-economic classification (1) | 59.9 (1441) | 54.0 (1641) | 38.5 (781) | 29.32 (275) | 51.1 (1712.1) | 61.5 (669) |  |
| Socio-economic classification (2) | 20.9 (502) | 21.1 (688) | 23.7 (463) | 15.41 (146) | 17.1 (574.1) | 22.9 (249) |  |
| Socio-economic classification (3) | 19.2 (358) | 24.8 (592) | 37.8 (569) | 26.05 (206) | 31.7 (1061.5) | 10.8 (117) |  |
| Managers, directors and senior SOC (1) | 9.6 (171) | 15.9 (434) | 11.2 (212) | 15.13 (112) | 11.0 (367.8) | NA |  |
| Professional SOC (2) | 31.8 (802) | 25.7 (812) | 16.3 (352) | 12.53 (92) | 22.4 (748.7) | NA |  |
| Associate professional and technical SOC (3) | 21.8 (547) | 14.1 (473) | 11.8 (235) | 14.41 (113) | 19.2 (642.8) | NA |  |
| Administrative and secretarial SOC (4) | 11.1 (256) | 12.4 (413) | 16.0 (317) | 16.74 (106) | 10.9 (363.3) | NA |  |
| Skilled trades SOC (5) | 4.6 (83) | 7.1 (163) | 9.5 (154) | 10.42 (64) | 8.3 (276.8) | NA |  |
| Caring, leisure and other service SOC (6) | 7.3 (181) | 8.0 (248) | 10.6 (205) | 9.14 (66) | 8.5 (283.6) | NA |  |
| Sales and customer service SOC (7) | 5.3 (124) | 4.4 (129) | 7.6 (118) | 4.96 (45) | 7.0 (234.8) | NA |  |
| Process, plant and machine operatives SOC (8) | 3.5 (55) | 6.3 (129) | 7.6 (104) | 7.39 (46) | 4.8 (160.4) | NA |  |
| Elementary occupations SOC (9) | 4.8 (82) | 6.1 (120) | 9.3 (116) | 9.27 (65) | 8.0 (269.5) | NA |  |
| Key worker | 61.3 (1293) | 58.9 (1742) | 62.2 (1127) | 48.06 (335) | 53.6 (1793.0) | 51.0 (554) |  |
| HH (Female, partner + children) | 12.5 (337) | 26.6 (911) | 8.5 (151) | 13.61 (111) | 14.7 (459.7) | 29.6 (322) |  |
| HH (Male, partner + children) | 15.4 (227) | 30.3 (788) | 12.3 (205) | 16.56 (87) | 17.9 (559.8) | 15.3 ( 166) |  |
| HH (Female, partner no children) | 17.1 (513) | 9.4 (334) | 21.7 (448) | 21.61 (189) | 19.2 (600.4) | 21.4 (233) |  |
| HH (Male, partner no children) | 13.8 (355) | 9.4 (239) | 26.3 (435) | 22.76 (156) | 19.9 (620.8) | 13.7 (149) |  |
| HH (lone parents) | 4.4 (86) | 8.0 (208) | 4.3 (79) | 5.2 (36) | 3.5 (110.5) | 6.1 (66) |  |
| HH (others, e.g., housemates) | 19.6 (429) | 2.8 (64) | 1.7 (25) | 3.39 (27) | 13.7 (428.3) | 1.7 (19) |  |
| HH (alone) | 17.0 (354) | 13.5 (377) | 25.0 (470) | 16.88 (103) | 11.0 (344.0) | 12.1 (132) |  |
| Overcrowding - Mean (SD) | 0.8 (0.5) | 0.6 (0.4) | 0.5 (0.3) | 0.50 (0.27) | 0.6 (0.3) | .45 (.21) |  |
| pre_Chronic health disability | 14.2 (380) | 30.7 (930) | 24.5 (486) | 13.8 (105) | 26.6 (889.5) | - |  |
| pre_Psychological Distress | 23.8 (562) | 14.2 (395) | 10.5 (193) | 8.04 (46) | 17.6 (588.2) | 8.9 (97) |  |
| pre_Low Life Satisfaction | 22.8 (498) | 12.0 (292) | 21.5 (390) | 24.46 (166) | 24.5 (818.6) | 12.8 (139) |  |
| pre_Fair/Poor Self-rated health | 7.9 (161) | 14.3 (368) | 12.2 (203) | 16.23 (85) | 13.8 (460.6) | - |  |
| pre_Often Lonely | 8.7 (215) | 5.8 (173) | 7.8 (124) | 4.28 (27) | 7.5 (251.1) | 8.6 (94) |  |
| pre_Less than daily Social Contact | 71.7 (1694) | 71.8 (2106) | 67.9 (1211) | 36.09 (239) | - | 33.0 (359) |  |
| **Total N** | 2301 | 2921 | 1813 | 709 | 3348 | 1087 |  |

Note: Values are % (n), unless otherwise specified.

N.B. For USOC, the descriptive statistics in Table S5 are based on data from a single wave at each timepoint (T1: May 2020; T2 September 2020; T3: January 2021), thus these samples differ from those used in the main analyses, which are based upon data from multiple waves.
