## Supplementary file 6 for "Home working and its association with social and mental wellbeing at different stages of the COVID-19 pandemic: Evidence from seven UK longitudinal population surveys"

**Supplementary file 6. Outcome Descriptives by Exposure**

| Outcome | Cohort | Exposure | Timepoint 1 | Timepoint 2 | Timepoint 3 |
| --- | --- | --- | --- | --- | --- |
| Psychological distress | Next steps | Employers | 33.6 | 26.2 | 31.6 |
|  |  | Home | 36.9 | 38.2 | 33.8 |
|  |  | Hybrid | - | 33.7 | 35.6 |
|  |  | Full sample | 35.9 | 32.1 | 33.3 |
|  | BCS70 | Employers | 15.4 | 21.5 | 17.8 |
|  |  | Home | 13.2 | 19.5 | 17.6 |
|  |  | Hybrid | - | 20.0 | 15.7 |
|  |  | Full sample | 14.1 | 20.8 | 17.3 |
|  | NCDS | Employers | 10.8 | 13.9 | 13.3 |
|  |  | Home | 9.6 | 8.3 | 11.9 |
|  |  | Hybrid | - | 18.0 | 13.6 |
|  |  | Full sample | 10.2 | 13.2 | 12.9 |
|  | ELSA | Employers | 18.3 | - | 21.9 |
|  |  | Home | 13.0 | - | 26.2 |
|  |  | Hybrid | - | - | 24.3 |
|  |  | Full sample | 15.9 | - | 23.3 |
|  | USOC | Employers | 23.1 | 21.2 | 25.2 |
|  |  | Home | 30.5 | 20.4 | 30.5 |
|  |  | Hybrid | - | 19.9 | 29.9 |
|  |  | Full sample | 26.9 | 20.7 | 28.0 |
| Low life satisfaction | Next steps | Employers | 29.5 | 27.9 | 37.6 |
|  |  | Home | 30.3 | 28.5 | 38.9 |
|  |  | Hybrid | - | 27.4 | 35.2 |
|  |  | Full sample | 30.1 | 28.0 | 37.7 |
|  | BCS70 | Employers | 24.9 | 26.2 | 30.4 |
|  |  | Home | 23.4 | 27.5 | 32.1 |
|  |  | Hybrid | - | 27.5 | 37.5 |
|  |  | Full sample | 24.0 | 26.8 | 32.3 |
|  | NCDS | Employers | 20.6 | 20.7 | 28.9 |
|  |  | Home | 21.1 | 23.1 | 33.5 |
|  |  | Hybrid | - | 26.9 | 27.1 |
|  |  | Full sample | 20.8 | 22.1 | 29.9 |
|  | ELSA | Employers | 24.7 | - | 33.1 |
|  |  | Home | 30.6 | - | 33.0 |
|  |  | Hybrid | - | - | 36.2 |
|  |  | Full sample | 27.4 | - | 33.6 |
|  | USOC | Employers | 35.2 | 32.6 | 36.6 |
|  |  | Home | 27.7 | 25.3 | 34.5 |
|  |  | Hybrid | - | 26.8 | 30.1 |
|  |  | Full sample | 31.9 | 29.6 | 34.7 |
| Poor self-rated health | Next steps | Employers | 4.5 | 7.4 | 8.5 |
|  |  | Home | 8.7 | 6.3 | 9.7 |
|  |  | Hybrid | - | 8.7 | 8.5 |
|  |  | Full sample | 7.4 | 7.2 | 9.0 |
|  | BCS70 | Employers | 7.4 | 8.5 | 14.6 |
|  |  | Home | 8.2 | 10.4 | 14.0 |
|  |  | Hybrid | - | 8.2 | 10.4 |
|  |  | Full sample | 7.8 | 8.9 | 13.7 |
|  | NCDS | Employers | 9.8 | 11.0 | 14.4 |
|  |  | Home | 8.7 | 6.3 | 12.7 |
|  |  | Hybrid | - | 8.2 | 8.2 |
|  |  | Full sample | 9.2 | 9.6 | 12.9 |
|  | ELSA | Employers | 24.8 | - | 21.2 |
|  |  | Home | 9.7 | - | 8.1 |
|  |  | Hybrid | - | - | 11.9 |
|  |  | Full sample | 17.9 | - | 16.6 |
|  | USOC | Employers | - | - | 11.9 |
|  |  | Home | - | - | 9.9 |
|  |  | Hybrid | - | - | 7.3 |
|  |  | Full sample | - | - | 10.4 |
| Often lonely | Next steps | Employers | 8.1 | 7.8 | 9.2 |
|  |  | Home | 9.9 | 6.8 | 11.4 |
|  |  | Hybrid | - | 7.2 | 9.2 |
|  |  | Full sample | 9.3 | 7.3 | 10.2 |
|  | BCS70 | Employers | 6.2 | 6.5 | 5.4 |
|  |  | Home | 4.0 | 6.6 | 5.8 |
|  |  | Hybrid | - | 5.4 | 4.6 |
|  |  | Full sample | 4.9 | 6.4 | 5.4 |
|  | NCDS | Employers | 4.2 | 3.8 | 4.0 |
|  |  | Home | 4.0 | 2.8 | 4.9 |
|  |  | Hybrid | - | 11.7 | 4.2 |
|  |  | Full sample | 4.1 | 4.7 | 4.3 |
|  | ELSA | Employers | 4.8 | - | 5.4 |
|  |  | Home | 2.6 | - | 9.5 |
|  |  | Hybrid | - | - | 2.4 |
|  |  | Full sample | 3.8 | - | 5.8 |
|  | USOC | Employers | 5.3 | 4.8 | 7.8 |
|  |  | Home | 10.6 | 5.3 | 9.7 |
|  |  | Hybrid | - | 3.3 | 6.4 |
|  |  | Full sample | 6.7 | 4.6 | 8.3 |
| Low social contact | Next steps | Employers | 68.8 | 69.3 | 75.0 |
|  |  | Home | 69.6 | 69.6 | 73.7 |
|  |  | Hybrid | - | 71.5 | 77.0 |
|  |  | Full sample | 69.3 | 69.8 | 74.8 |
|  | BCS70 | Employers | 67.8 | 69.3 | 70.9 |
|  |  | Home | 64.5 | 73.5 | 74.8 |
|  |  | Hybrid | - | 73.4 | 75.7 |
|  |  | Full sample | 65.8 | 71.0 | 73.3 |
|  | NCDS | Employers | 60.7 | 63.7 | 65.9 |
|  |  | Home | 62.2 | 68.3 | 71.2 |
|  |  | Hybrid | - | 69.9 | 70.6 |
|  |  | Full sample | 61.4 | 65.6 | 68.0 |
|  | ELSA | Employers | 48.1 | - | 49.3 |
|  |  | Home | 41.3 | - | 50.2 |
|  |  | Hybrid | - | - | 57.4 |
|  |  | Full sample | 45.0 | - | 50.9 |
|  | USOC* | Employers | 57.5 | - | 53.9 |
|  |  | Home | 51.8 | - | 48.5 |
|  |  | Hybrid | - | - | 50.6 |
|  |  | Full sample | 54.4 | - | 51.7 |

N.B. For USOC, the descriptive statistics in Table 1 are based on data from a single wave (T1: May 2020; T2 September 2020; T3: January 2021), thus the samples used to derive these descriptives differ from those used in the main analyses, which are based upon data from multiple waves.

*Descriptive statistics based on data from June 2020 (T1) and November 2020 (T2) as these are the only waves at which information on social contact were collected in USOC.
