## Supplementary file 7 for "Home working and its association with social and mental wellbeing at different stages of the COVID-19 pandemic: Evidence from seven UK longitudinal population surveys"

**Supplementary file 7. Pooled analyses for the main results (Forest plots)**

### No adjustment

#### Low life satisfaction

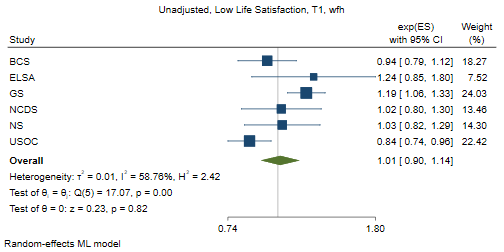

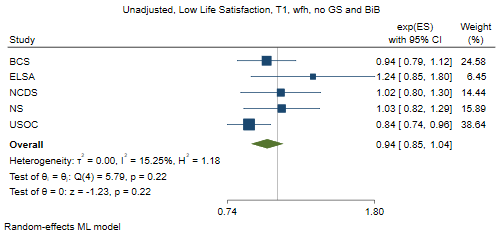

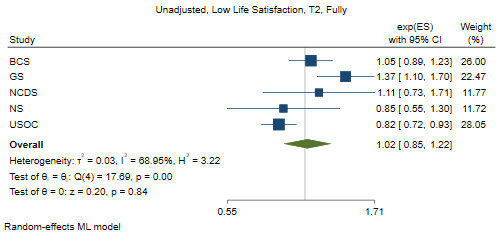

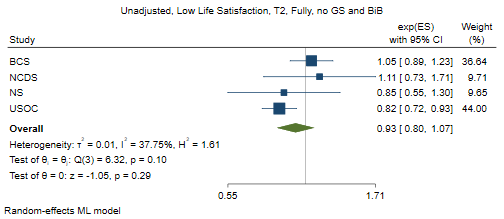

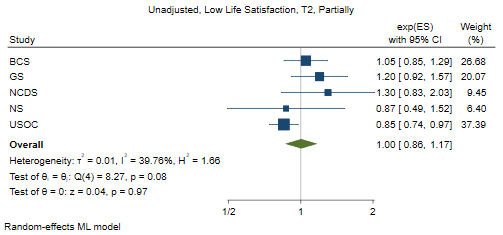

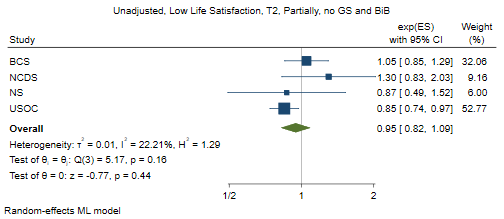

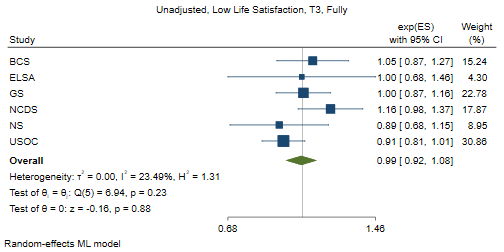

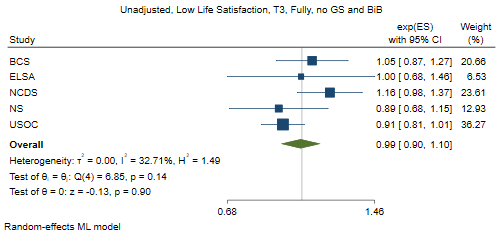

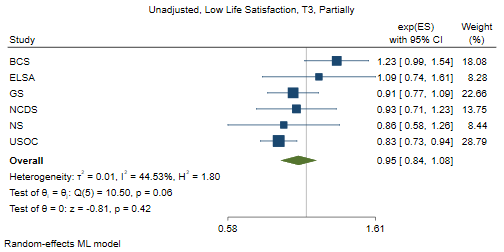

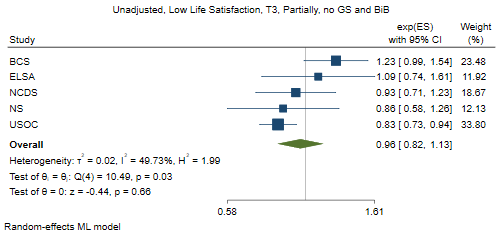

#### Often lonely

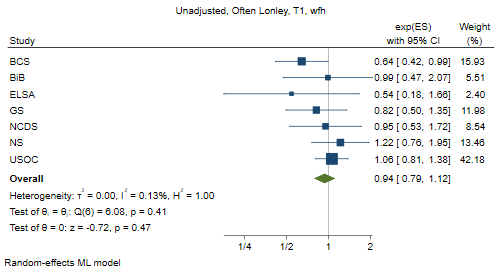

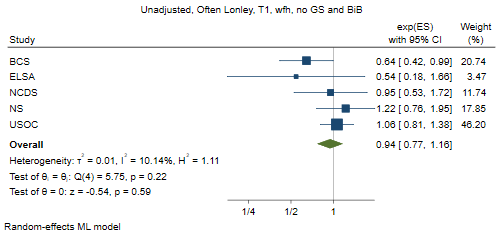

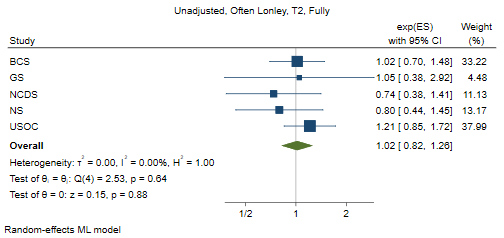

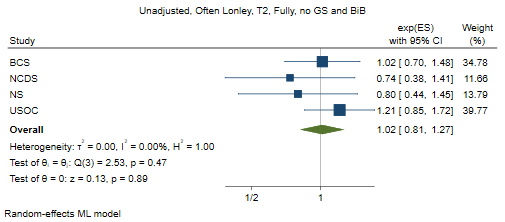

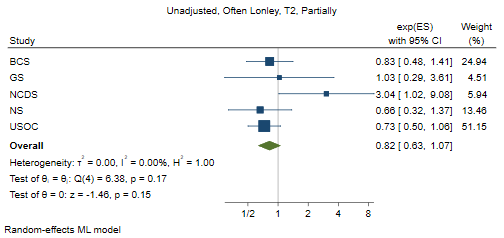

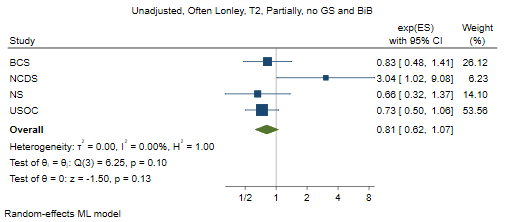

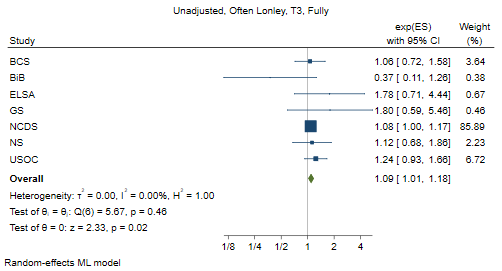

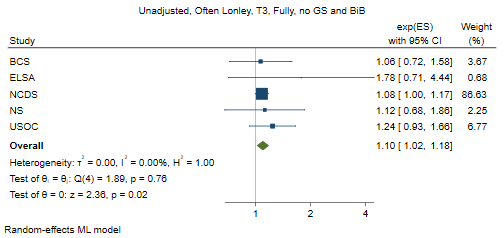

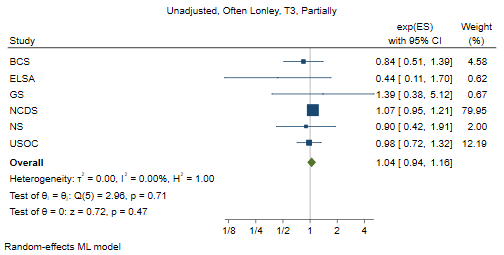

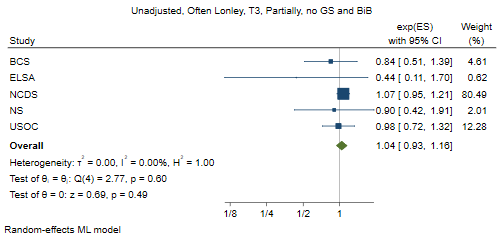

#### Psychological distress

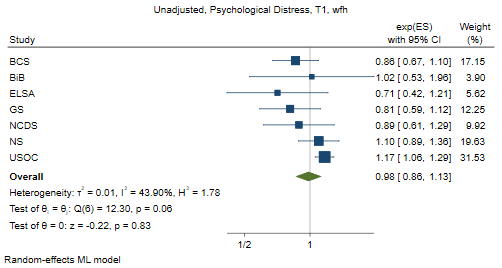

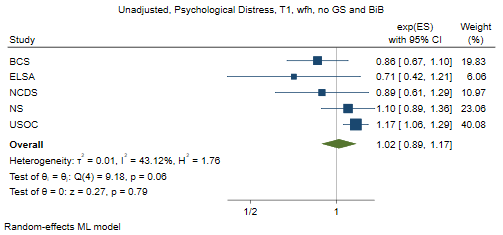

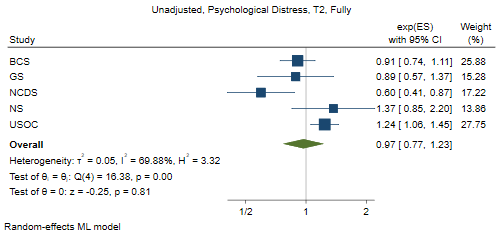

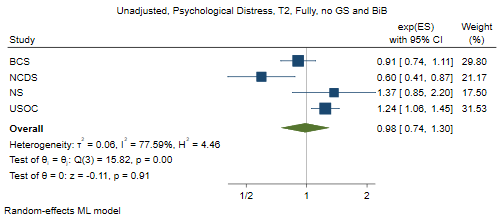

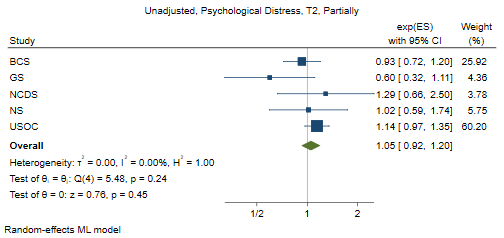

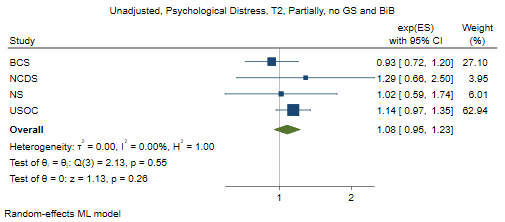

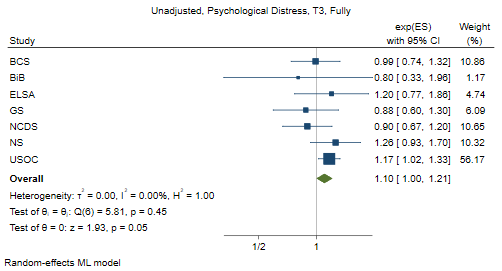

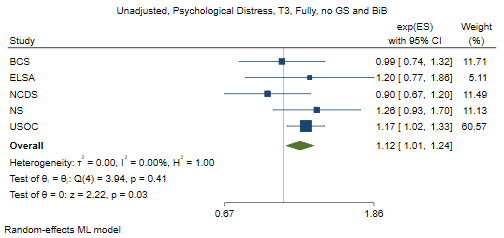

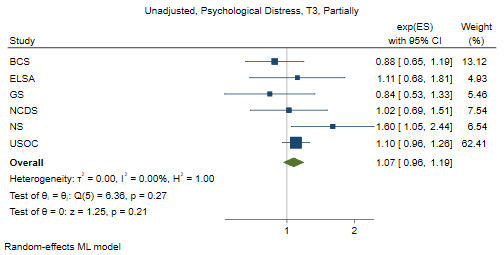

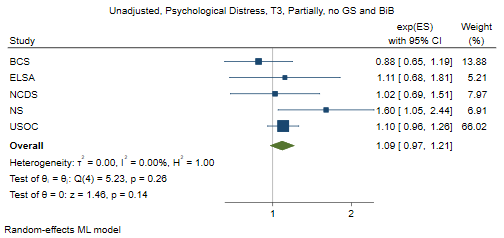

#### Poor self-rated health

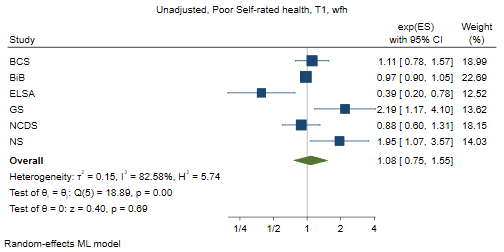

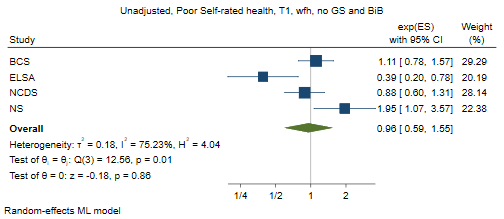

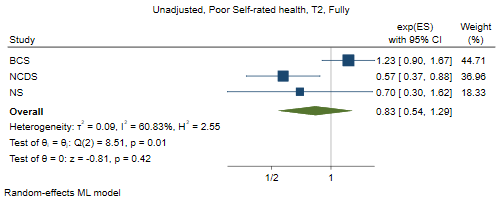

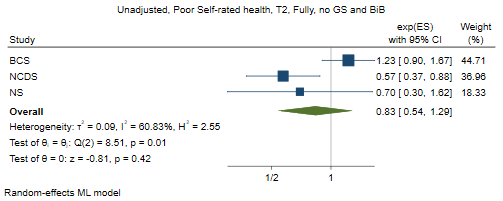

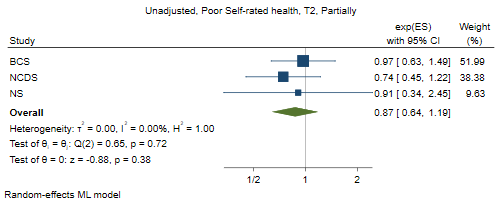

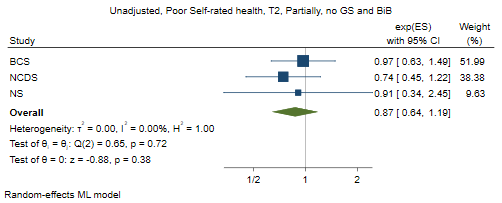

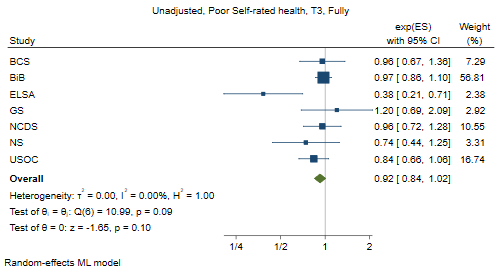

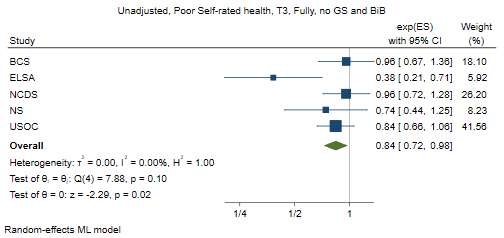

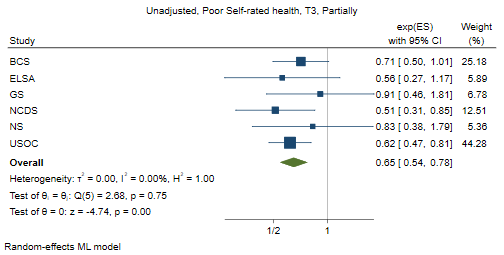

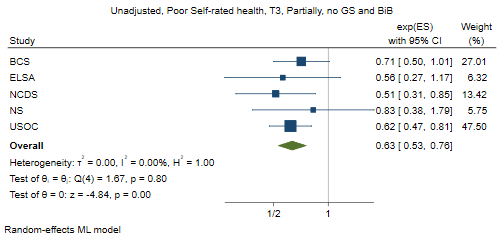

#### Low social contact

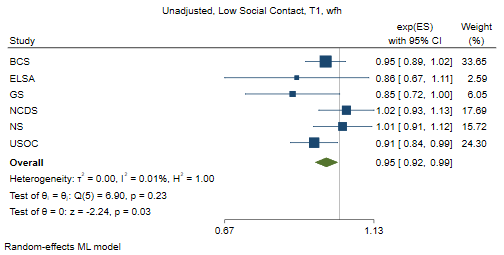

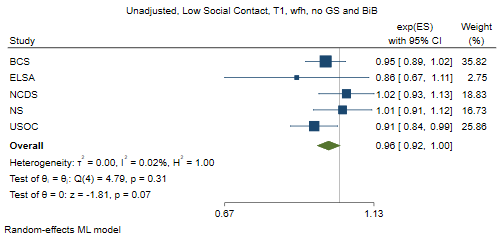

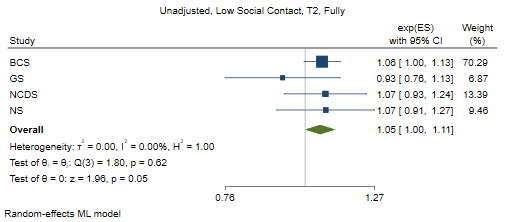

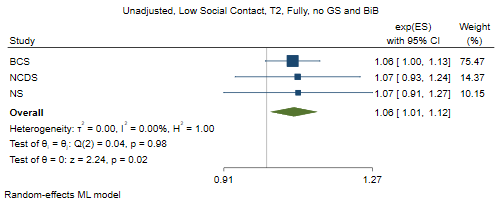

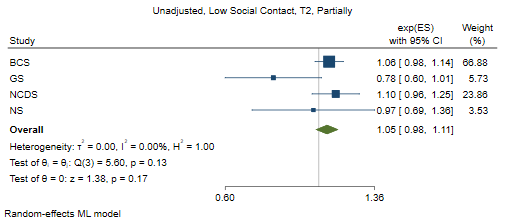

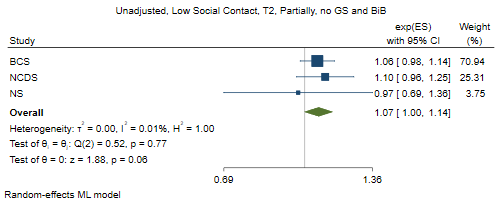

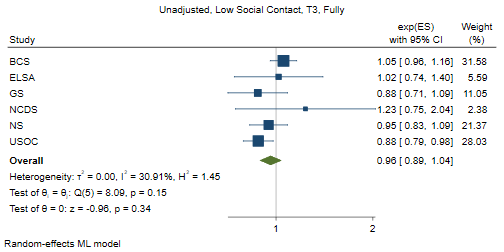

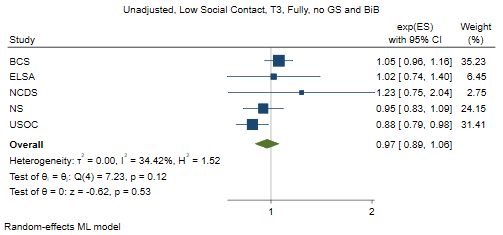

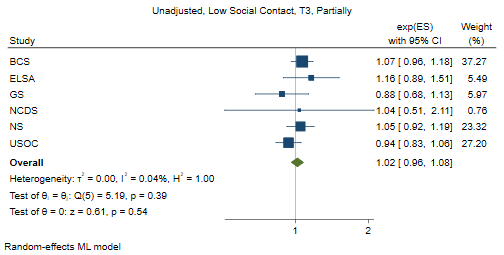

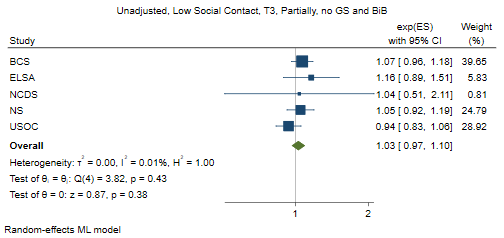

### Socio-demographic adjustment

#### Low life satisfaction

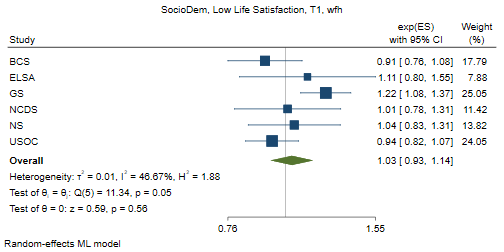

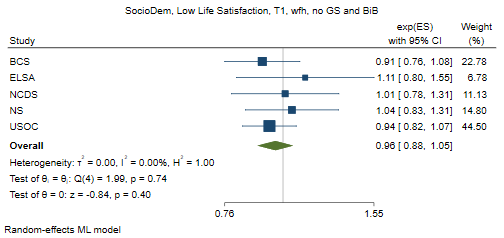

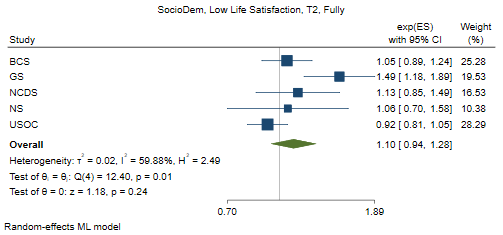

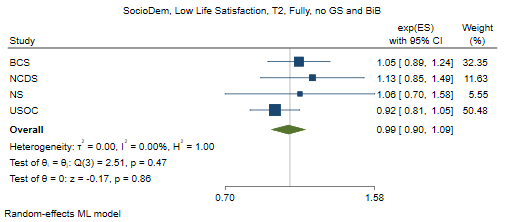

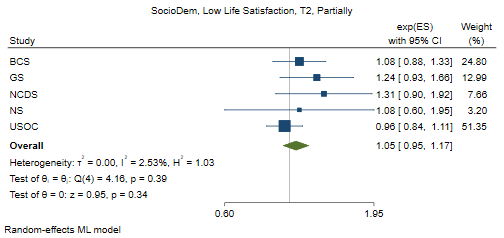

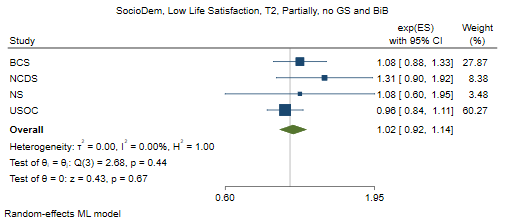

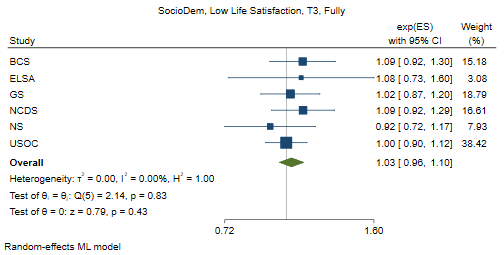

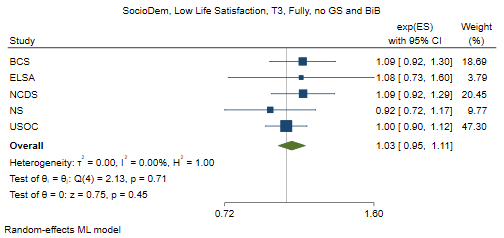

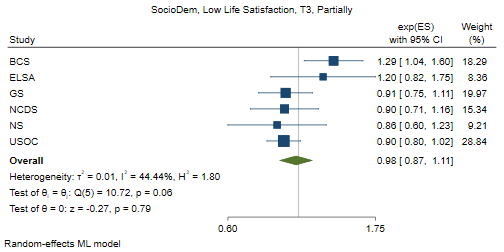

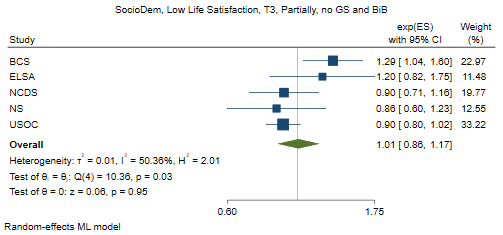

#### Often lonely

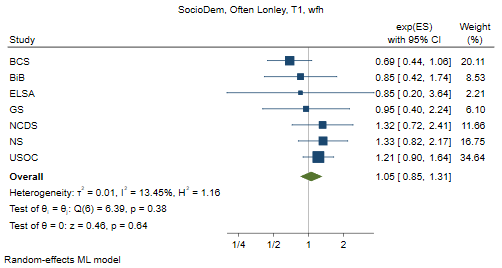

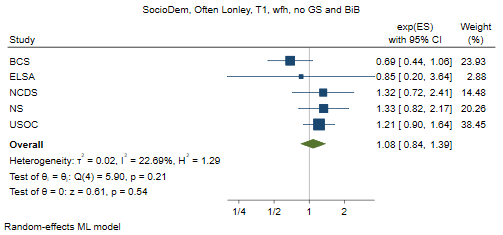

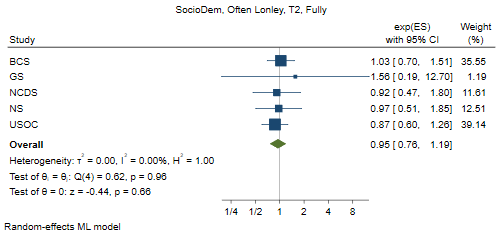

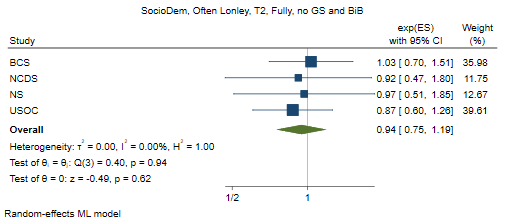

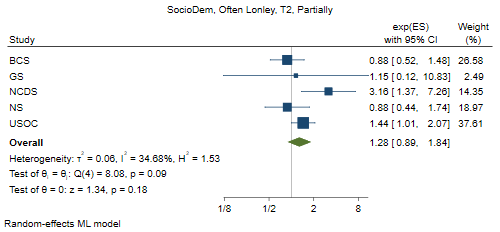

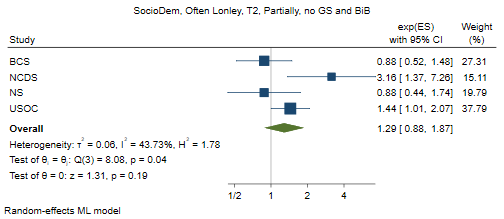

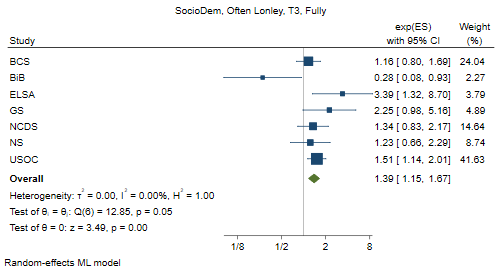

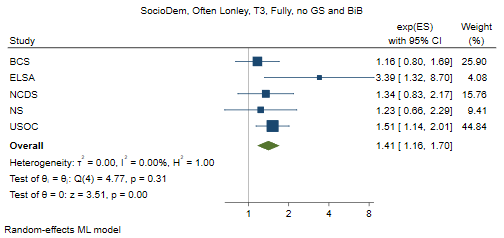

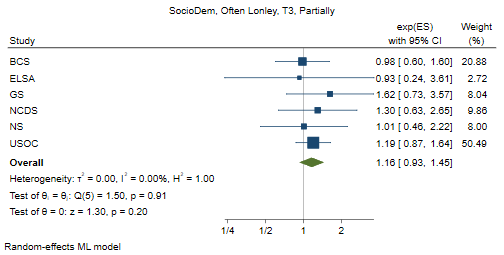

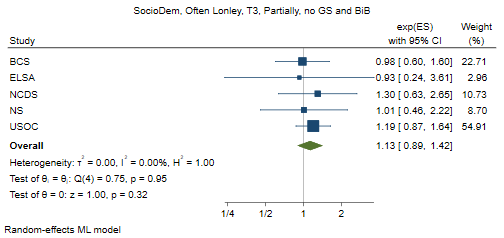

#### Psychological distress

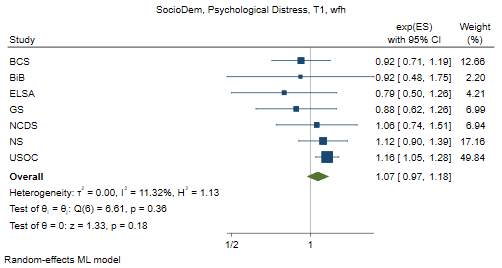

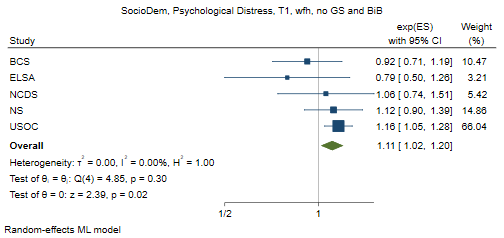

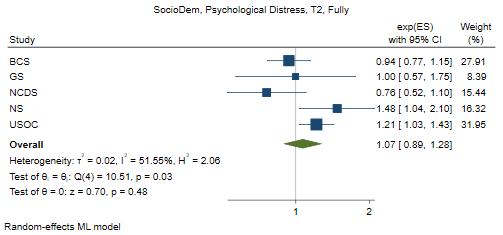

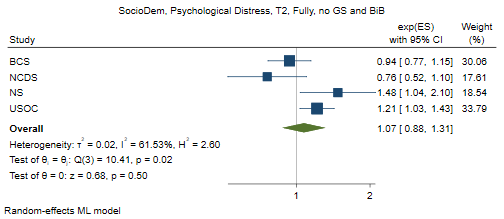

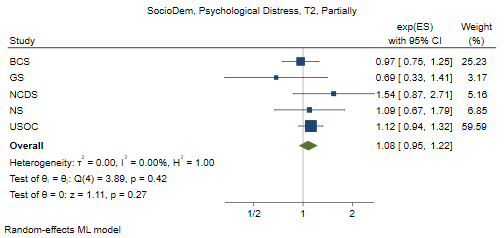

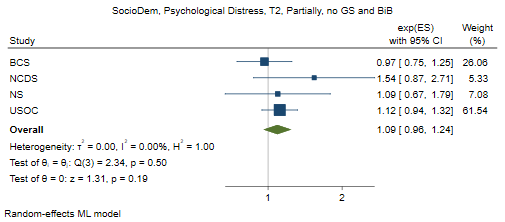

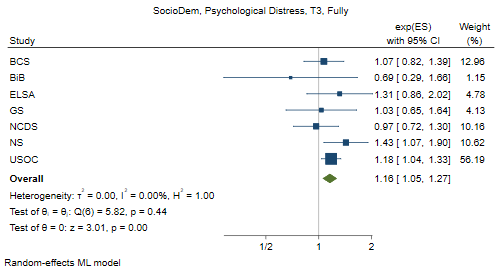

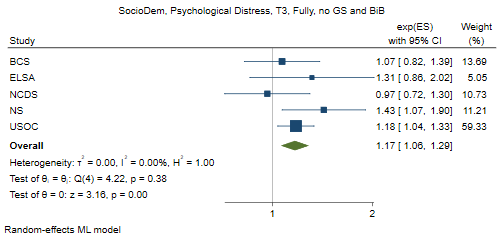

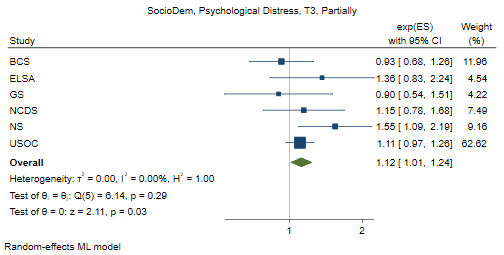

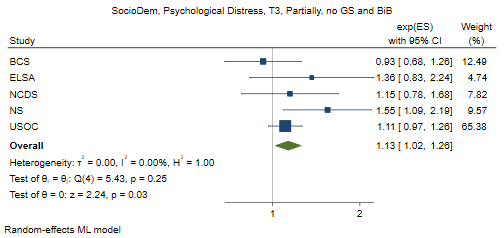

#### Poor self-rated health

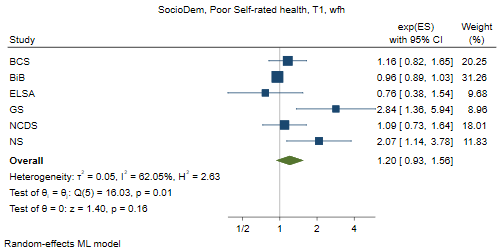

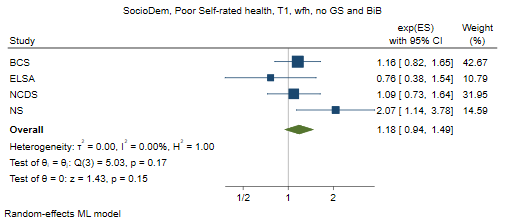

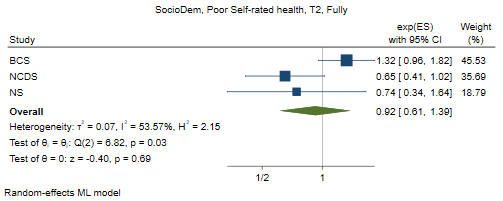

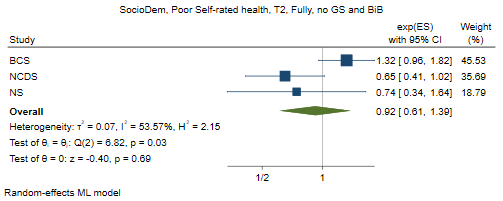

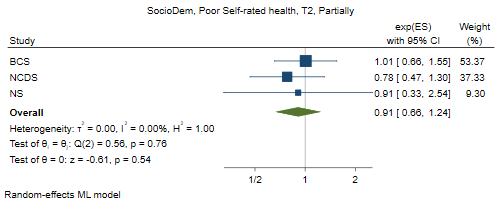

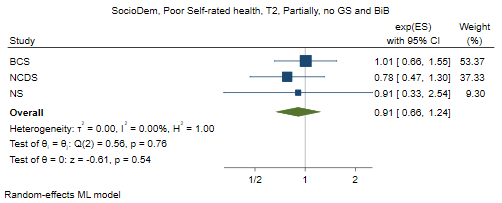

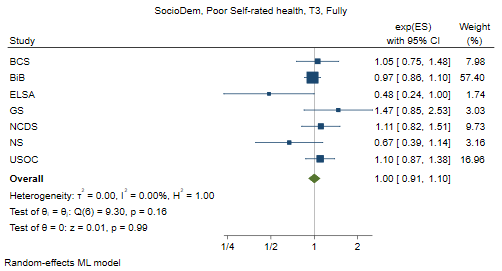

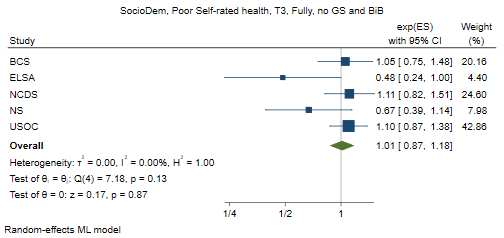

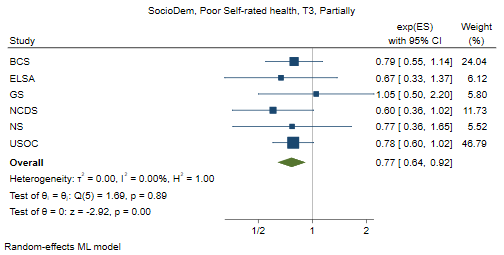

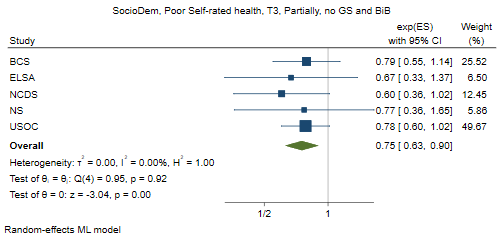

#### Low social contact

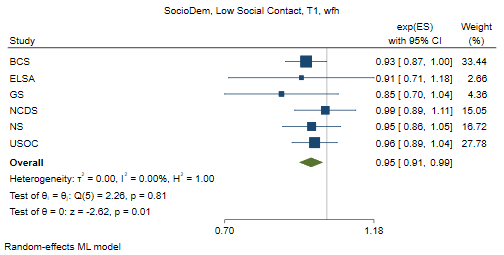

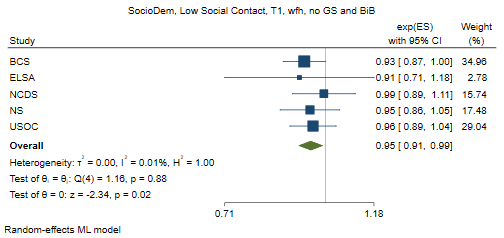

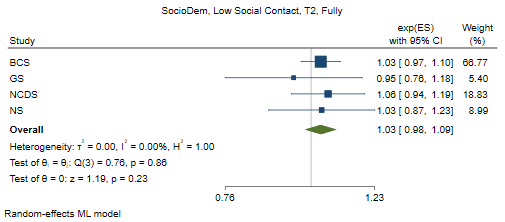

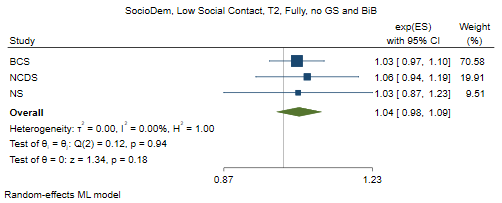

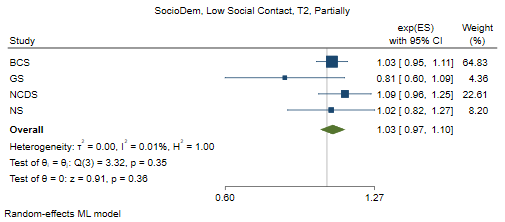

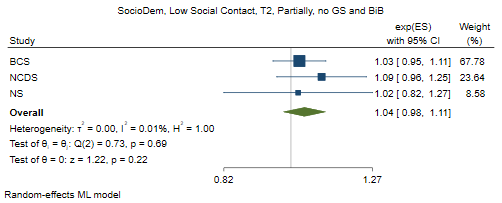

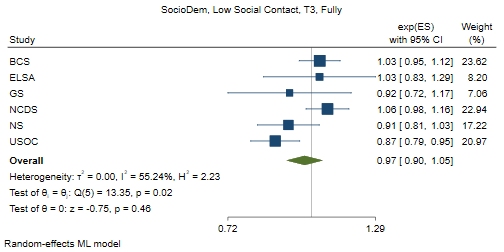

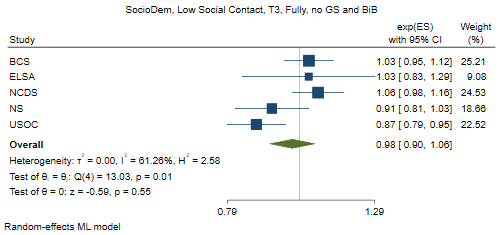

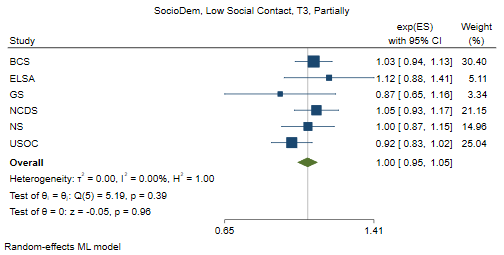

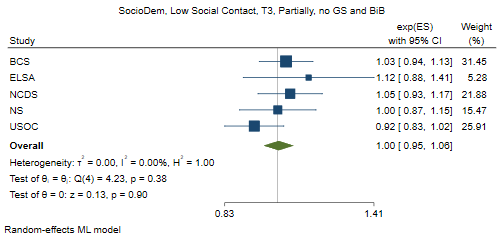

### Job adjustment

#### Low life satisfaction

#### Often lonely

#### Psychological distress

#### Poor self-rated health

w

#### social contact

### Full adjustment

#### Low life satisfaction

#### Often lonely

#### Psychological distress

#### Poor self-rated health

#### Low social contact
