## Supplementary file 8 for "Home working and its association with social and mental wellbeing at different stages of the COVID-19 pandemic: Evidence from seven UK longitudinal population surveys"

**Supplementary file 8. Results from subgroup meta-analysis for between-group heterogeneity tests**

|  | Outcome | Exposure | Sex (male, female) | Education (university degree or below) | Age group (16-29, 30-49, 50+) | Hours (full time, part time) |
| --- | --- | --- | --- | --- | --- | --- |
| Wave 1 | 1 LifeSatBin | WFH | Q_b = chi2(1) = 0.53 Prob > Q_b = 0.468 | Q_b = chi2(1) = 0.15 Prob > Q_b = 0.696 | Q_b = chi2(2) = 0.80 Prob > Q_b = 0.672 | Q_b = chi2(1) = 0.40 Prob > Q_b = 0.526 |
|  | 2 LonBin | WFH | Q_b = chi2(1) = 0.16 Prob > Q_b = 0.689 | Q_b = chi2(1) = 2.80 Prob > Q_b = 0.094 | **Q_b = chi2(2) = 6.46 Prob > Q_b = 0.040** | Q_b = chi2(1) = 1.50 Prob > Q_b = 0.220 |
|  | 3 PsyDisBin | WFH | Q_b = chi2(1) = 0.56 Prob > Q_b = 0.454 | Q_b = chi2(1) = 2.01 Prob > Q_b = 0.156 | Q_b = chi2(2) = 3.00 Prob > Q_b = 0.223 | Q_b = chi2(1) = 0.09 Prob > Q_b = 0.770 |
|  | 4 SRHBin | WFH | Q_b = chi2(1) = 0.00 Prob > Q_b = 0.962 | Q_b = chi2(1) = 1.16 Prob > Q_b = 0.282 | **Q_b = chi2(1) = 5.09 Prob > Q_b = 0.024** | Q_b = chi2(1) = 0.22 Prob > Q_b = 0.638 |
|  | 5 SocCont | WFH | Q_b = chi2(1) = 1.91 Prob > Q_b = 0.167 | Q_b = chi2(1) = 0.41 Prob > Q_b = 0.522 | Q_b = chi2(2) = 0.36 Prob > Q_b = 0.835 | Q_b = chi2(1) = 2.79 Prob > Q_b = 0.095 |
| Wave 2 | 1 LifeSatBin | Fully WFH | Q_b = chi2(1) = 0.17 Prob > Q_b = 0.677 | Q_b = chi2(1) = 0.33 Prob > Q_b = 0.568 | Q_b = chi2(2) = 3.51 Prob > Q_b = 0.173 | Q_b = chi2(1) = 0.20 Prob > Q_b = 0.653 |
|  | 2 LonBin | Fully WFH | Q_b = chi2(1) = 0.52 Prob > Q_b = 0.469 | Q_b = chi2(1) = 1.16 Prob > Q_b = 0.281 | Q_b = chi2(2) = 3.38 Prob > Q_b = 0.184 | Q_b = chi2(1) = 0.11 Prob > Q_b = 0.746 |
|  | 3 PsyDisBin | Fully WFH | Q_b = chi2(1) = 0.32 Prob > Q_b = 0.572 | Q_b = chi2(1) = 0.00 Prob > Q_b = 0.966 | Q_b = chi2(2) = 0.45 Prob > Q_b = 0.797 | Q_b = chi2(1) = 0.98 Prob > Q_b = 0.323 |
|  | 4 SRHBin | Fully WFH | Q_b = chi2(1) = 0.16 Prob > Q_b = 0.692 | Q_b = chi2(1) = 0.07 Prob > Q_b = 0.791 | Q_b = chi2(1) = 0.51 Prob > Q_b = 0.476 | Q_b = chi2(1) = 0.06 Prob > Q_b = 0.810 |
|  | 5 SocCont | Fully WFH | Q_b = chi2(1) = 1.09 Prob > Q_b = 0.297 | Q_b = chi2(1) = 1.16 Prob > Q_b = 0.281 | Q_b = chi2(1) = 0.01 Prob > Q_b = 0.939 | Q_b = chi2(1) = 0.01 Prob > Q_b = 0.910 |
|  | 1 LifeSatBin | Partially WFH | Q_b = chi2(1) = 1.73 Prob > Q_b = 0.189 | **Q_b = chi2(1) = 4.39 Prob > Q_b = 0.036** | Q_b = chi2(2) = 4.06 Prob > Q_b = 0.131 | Q_b = chi2(1) = 0.80 Prob > Q_b = 0.372 |
|  | 2 LonBin | Partially WFH | **Q_b = chi2(1) = 4.96 Prob > Q_b = 0.026** | Q_b = chi2(1) = 1.71 Prob > Q_b = 0.191 | Q_b = chi2(2) = 2.23 Prob > Q_b = 0.327 | Q_b = chi2(1) = 1.90 Prob > Q_b = 0.168 |
|  | 3 PsyDisBin | Partially WFH | Q_b = chi2(1) = 0.16 Prob > Q_b = 0.690 | Q_b = chi2(1) = 2.69 Prob > Q_b = 0.101 | Q_b = chi2(2) = 5.64 Prob > Q_b = 0.060 | **Q_b = chi2(1) = 13.24 Prob > Q_b = 0.000** |
|  | 4 SRHBin | Partially WFH | **Q_b = chi2(1) = 15.75 Prob > Q_b = 0.000** | Q_b = chi2(1) = 0.07 Prob > Q_b = 0.788 | Q_b = chi2(1) = 0.99 Prob > Q_b = 0.320 | Q_b = chi2(1) = 1.99 Prob > Q_b = 0.159 |
|  | 5 SocCont | Partially WFH | Q_b = chi2(1) = 0.06 Prob > Q_b = 0.812 | Q_b = chi2(1) = 0.00 Prob > Q_b = 0.967 | Q_b = chi2(1) = 0.01 Prob > Q_b = 0.940 | Q_b = chi2(1) = 0.90 Prob > Q_b = 0.342 |
| Wave 3 | 1 LifeSatBin | Fully WFH | Q_b = chi2(1) = 0.04 Prob > Q_b = 0.850 | Q_b = chi2(1) = 2.10 Prob > Q_b = 0.147 | Q_b = chi2(2) = 1.64 Prob > Q_b = 0.441 | Q_b = chi2(1) = 0.54 Prob > Q_b = 0.463 |
|  | 2 LonBin | Fully WFH | Q_b = chi2(1) = 0.70 Prob > Q_b = 0.401 | Q_b = chi2(1) = 2.56 Prob > Q_b = 0.110 | Q_b = chi2(2) = 0.72 Prob > Q_b = 0.697 | Q_b = chi2(1) = 0.18 Prob > Q_b = 0.675 |
|  | 3 PsyDisBin | Fully WFH | Q_b = chi2(1) = 0.08 Prob > Q_b = 0.774 | Q_b = chi2(1) = 0.60 Prob > Q_b = 0.437 | Q_b = chi2(2) = 0.57 Prob > Q_b = 0.753 | **Q_b = chi2(1) = 7.13 Prob > Q_b = 0.008** |
|  | 4 SRHBin | Fully WFH | Q_b = chi2(1) = 0.18 Prob > Q_b = 0.668 | Q_b = chi2(1) = 0.00 Prob > Q_b = 0.969 | Q_b = chi2(2) = 0.28 Prob > Q_b = 0.870 | Q_b = chi2(1) = 0.06 Prob > Q_b = 0.801 |
|  | 5 SocCont | Fully WFH | Q_b = chi2(1) = 1.85 Prob > Q_b = 0.174 | Q_b = chi2(1) = 0.16 Prob > Q_b = 0.685 | **Q_b = chi2(2) = 6.66 Prob > Q_b = 0.036** | Q_b = chi2(1) = 0.79 Prob > Q_b = 0.373 |
|  | 1 LifeSatBin | Partially WFH | Q_b = chi2(1) = 0.01 Prob > Q_b = 0.938 | Q_b = chi2(1) = 0.30 Prob > Q_b = 0.582 | Q_b = chi2(2) = 1.49 Prob > Q_b = 0.475 | Q_b = chi2(1) = 1.87 Prob > Q_b = 0.171 |
|  | 2 LonBin | Partially WFH | Q_b = chi2(1) = 0.75 Prob > Q_b = 0.386 | **Q_b = chi2(1) = 3.83 Prob > Q_b = 0.050** | Q_b = chi2(2) = 0.80 Prob > Q_b = 0.672 | Q_b = chi2(1) = 0.25 Prob > Q_b = 0.619 |
|  | 3 PsyDisBin | Partially WFH | Q_b = chi2(1) = 1.11 Prob > Q_b = 0.292 | **Q_b = chi2(1) = 6.12 Prob > Q_b = 0.013** | Q_b = chi2(2) = 5.31 Prob > Q_b = 0.070 | Q_b = chi2(1) = 0.00 Prob > Q_b = 0.970 |
|  | 4 SRHBin | Partially WFH | Q_b = chi2(1) = 1.46 Prob > Q_b = 0.227 | Q_b = chi2(1) = 0.08 Prob > Q_b = 0.775 | Q_b = chi2(2) = 1.73 Prob > Q_b = 0.421 | **Q_b = chi2(1) = 4.34 Prob > Q_b = 0.037** |
|  | 5 SocCont | Partially WFH | Q_b = chi2(1) = 1.36 Prob > Q_b = 0.244 | **Q_b = chi2(1) = 4.60 Prob > Q_b = 0.032** | Q_b = chi2(2) = 4.55 Prob > Q_b = 0.103 | Q_b = chi2(1) = 0.88 Prob > Q_b = 0.348 |
