## Supplementary file 9 for "Home working and its association with social and mental wellbeing at different stages of the COVID-19 pandemic: Evidence from seven UK longitudinal population surveys"

**Supplementary file 9. Sensitivity analyses of self-reported vs. imputed pre-pandemic home working (Usoc)**

| **TABLE 1) MODELS ADJUSTED FOR BASELINE WORKING FROM HOME** | | | | | |  |  |  |  |
| --- | --- | --- | --- | --- | --- | --- | --- | --- | --- |
| cohort | outcome | wave | exposure | adjustments | Raw coef (not exp) | coef_se | lower_ci | upper_ci | n |
| USOC | PsyDisBin | T1 | fully wfh | full | 0.06924 | 0.05303 | -0.03 | 0.17 | 14013 |
| USOC | PsyDisBin | T2 | fully wfh | full | 0.02563 | 0.09224 | -0.16 | 0.21 | 8573 |
| USOC | PsyDisBin | T2 | partially wfh | full | 0.01298 | 0.08713 | -0.16 | 0.18 | 8573 |
| USOC | PsyDisBin | T3 | fully wfh | full | 0.09054 | 0.06943 | -0.05 | 0.23 | 11484 |
| USOC | PsyDisBin | T3 | partially wfh | full | 0.11034 | 0.07003 | -0.03 | 0.25 | 11484 |
| USOC | LifeSatBin | T1 | fully wfh | full | -0.03212 | 0.06908 | -0.17 | 0.10 | 4462 |
| USOC | LifeSatBin | T2 | fully wfh | full | -0.14163 | 0.07282 | -0.28 | 0.00 | 8573 |
| USOC | LifeSatBin | T2 | partially wfh | full | -0.07149 | 0.07006 | -0.21 | 0.07 | 8573 |
| USOC | LifeSatBin | T3 | fully wfh | full | 0.01074 | 0.06379 | -0.11 | 0.14 | 11484 |
| USOC | LifeSatBin | T3 | partially wfh | full | -0.08086 | 0.06318 | -0.20 | 0.04 | 11484 |
| USOC | SRHBin | T1 | fully wfh | full |  |  |  |  |  |
| USOC | SRHBin | T2 | fully wfh | full |  |  |  |  |  |
| USOC | SRHBin | T2 | partially wfh | full |  |  |  |  |  |
| USOC | SRHBin | T3 | fully wfh | full | 0.07468 | 0.1211 | -0.16 | 0.31 | 11484 |
| USOC | SRHBin | T3 | partially wfh | full | -0.11251 | 0.12529 | -0.36 | 0.13 | 11484 |
| USOC | LonBin | T1 | fully wfh | full | 0.17852 | 0.157 | -0.13 | 0.49 | 14013 |
| USOC | LonBin | T2 | fully wfh | full | 0.08338 | 0.20166 | -0.31 | 0.48 | 8573 |
| USOC | LonBin | T2 | partially wfh | full | -0.24022 | 0.18541 | -0.60 | 0.12 | 8573 |
| USOC | LonBin | T3 | fully wfh | full | 0.38782 | 0.16932 | 0.06 | 0.72 | 11484 |
| USOC | LonBin | T3 | partially wfh | full | 0.22787 | 0.17355 | -0.11 | 0.57 | 11484 |
| USOC | SocCont | T1 | fully wfh | full | -0.03848 | 0.04404 | -0.12 | 0.05 | 4319 |
| USOC | SocCont | T2 | fully wfh | full |  |  |  |  |  |
| USOC | SocCont | T2 | partially wfh | full |  |  |  |  |  |
| USOC | SocCont | T3 | fully wfh | full | -0.1098 | 0.06346 | -0.23 | 0.01 | 3985 |
| USOC | SocCont | T3 | partially wfh | full | -0.06647 | 0.05939 | -0.18 | 0.05 | 3985 |

| **TABLE 2) MODELS ADJUSTED FOR PREDICTED PROBABILITY OF WORKING FROM HOME (PLUS SOC TYPE)** | | | | | | | | |  |
| --- | --- | --- | --- | --- | --- | --- | --- | --- | --- |
| cohort | outcome | wave | exposure | adjustments | Raw coef (not exp) | coef_se | lower_ci | upper_ci | n |
| USOC | PsyDisBin | T1 | fully wfh | full | 0.072925 | 0.051623 | -0.03 | 0.17 | 14013 |
| USOC | PsyDisBin | T2 | fully wfh | full | 0.032185 | 0.090121 | -0.14 | 0.21 | 8573 |
| USOC | PsyDisBin | T2 | partially wfh | full | 0.01315 | 0.084868 | -0.15 | 0.18 | 8573 |
| USOC | PsyDisBin | T3 | fully wfh | full | 0.106047 | 0.067702 | -0.03 | 0.24 | 11484 |
| USOC | PsyDisBin | T3 | partially wfh | full | 0.090003 | 0.068004 | -0.04 | 0.22 | 11484 |
| USOC | LifeSatBin | T1 | fully wfh | full | -0.03269 | 0.067193 | -0.16 | 0.10 | 4462 |
| USOC | LifeSatBin | T2 | fully wfh | full | -0.09915 | 0.069919 | -0.24 | 0.04 | 8573 |
| USOC | LifeSatBin | T2 | partially wfh | full | -0.05171 | 0.067346 | -0.18 | 0.08 | 8573 |
| USOC | LifeSatBin | T3 | fully wfh | full | -0.00697 | 0.060112 | -0.12 | 0.11 | 11484 |
| USOC | LifeSatBin | T3 | partially wfh | full | -0.09181 | 0.061509 | -0.21 | 0.03 | 11484 |
| USOC | SRHBin | T1 | fully wfh | full |  |  |  |  |  |
| USOC | SRHBin | T2 | fully wfh | full |  |  |  |  |  |
| USOC | SRHBin | T2 | partially wfh | full |  |  |  |  |  |
| USOC | SRHBin | T3 | fully wfh | full | 0.075238 | 0.116059 | -0.15 | 0.30 | 11484 |
| USOC | SRHBin | T3 | partially wfh | full | -0.11021 | 0.125392 | -0.36 | 0.14 | 11484 |
| USOC | LonBin | T1 | fully wfh | full | 0.14047 | 0.161588 | -0.18 | 0.46 | 14013 |
| USOC | LonBin | T2 | fully wfh | full | 0.108048 | 0.19533 | -0.27 | 0.49 | 8573 |
| USOC | LonBin | T2 | partially wfh | full | -0.23815 | 0.184703 | -0.60 | 0.12 | 8573 |
| USOC | LonBin | T3 | fully wfh | full | 0.394329 | 0.166246 | 0.07 | 0.72 | 11484 |
| USOC | LonBin | T3 | partially wfh | full | 0.227128 | 0.171957 | -0.11 | 0.56 | 11484 |
| USOC | SocCont | T1 | fully wfh | full | -0.02816 | 0.042937 | -0.11 | 0.06 | 4319 |
| USOC | SocCont | T2 | fully wfh | full |  |  |  |  |  |
| USOC | SocCont | T2 | partially wfh | full |  |  |  |  |  |
| USOC | SocCont | T3 | fully wfh | full | -0.17910 | 0.05638 | -0.29 | -0.07 | 3985 |
| USOC | SocCont | T3 | partially wfh | full | -0.09653 | 0.05651 | -0.21 | 0.01 | 3985 |

| **Table 3) Difference in estimates** | | |  |  |  |
| --- | --- | --- | --- | --- | --- |
| cohort | outcome | wave | exposure | adjustments | Table 1 minus Table 2 |
| USOC | PsyDisBin | T1 | fully wfh | full | -0.0037 |
| USOC | PsyDisBin | T2 | fully wfh | full | -0.0066 |
| USOC | PsyDisBin | T2 | partially wfh | full | -0.0002 |
| USOC | PsyDisBin | T3 | fully wfh | full | -0.0155 |
| USOC | PsyDisBin | T3 | partially wfh | full | 0.0203 |
| USOC | LifeSatBin | T1 | fully wfh | full | 0.0006 |
| USOC | LifeSatBin | T2 | fully wfh | full | -0.0425 |
| USOC | LifeSatBin | T2 | partially wfh | full | -0.0198 |
| USOC | LifeSatBin | T3 | fully wfh | full | 0.0177 |
| USOC | LifeSatBin | T3 | partially wfh | full | 0.0110 |
| USOC | SRHBin | T1 | fully wfh | full |  |
| USOC | SRHBin | T2 | fully wfh | full |  |
| USOC | SRHBin | T2 | partially wfh | full |  |
| USOC | SRHBin | T3 | fully wfh | full | -0.0006 |
| USOC | SRHBin | T3 | partially wfh | full | -0.0023 |
| USOC | LonBin | T1 | fully wfh | full | 0.0381 |
| USOC | LonBin | T2 | fully wfh | full | -0.0247 |
| USOC | LonBin | T2 | partially wfh | full | -0.0021 |
| USOC | LonBin | T3 | fully wfh | full | -0.0065 |
| USOC | LonBin | T3 | partially wfh | full | 0.0007 |
| USOC | SocCont | T1 | fully wfh | full | -0.0103 |
| USOC | SocCont | T2 | fully wfh | full |  |
| USOC | SocCont | T2 | partially wfh | full |  |
| USOC | SocCont | T3 | fully wfh | full | 0.0693 |
| USOC | SocCont | T3 | partially wfh | full | 0.0301 |
