## Supplementary figures and images for "Home working and its association with social and mental wellbeing at different stages of the COVID-19 pandemic: Evidence from seven UK longitudinal population surveys"

### Supplementary file 10

**Supplementary file 10. Main estimates (excluding BiB and GS)**

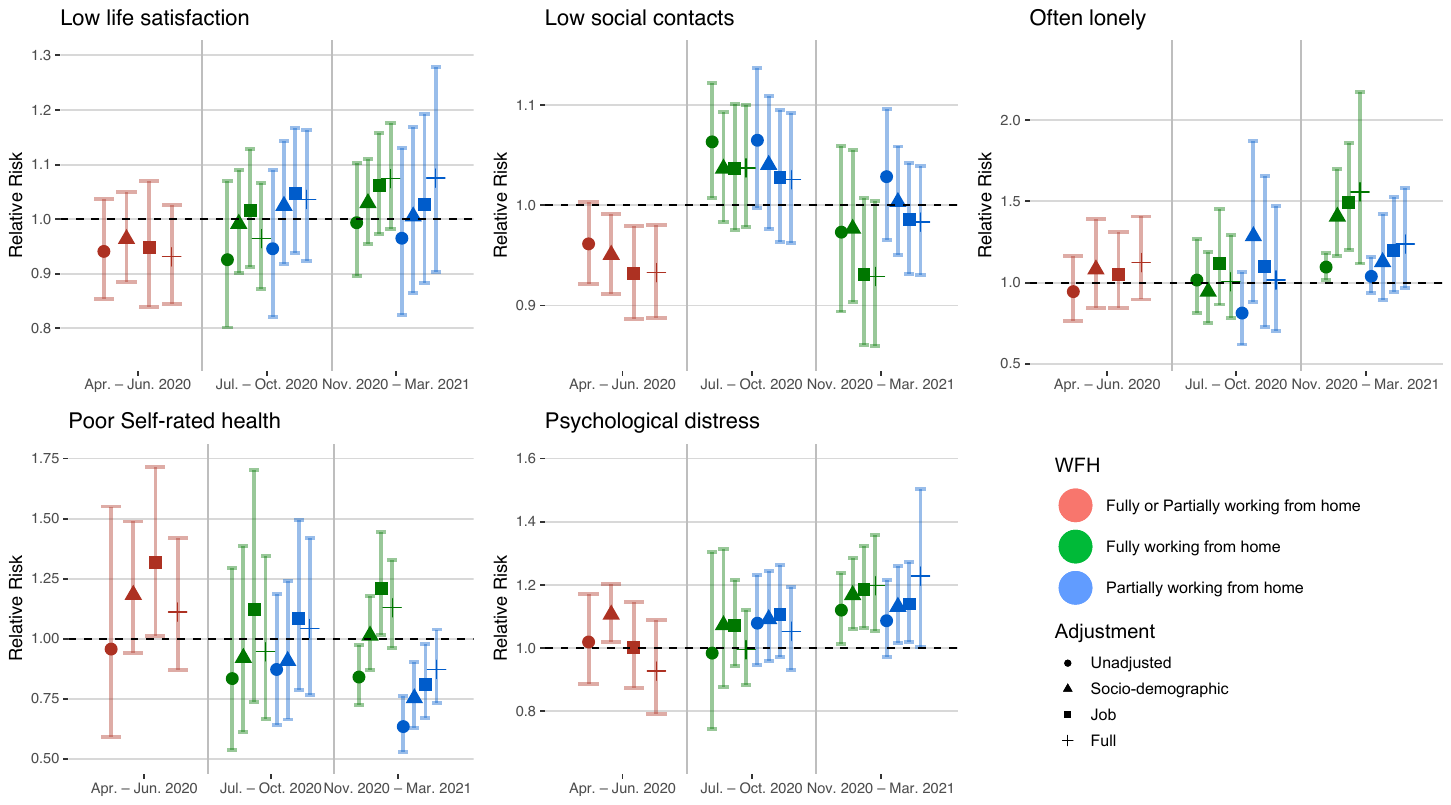
